# Sex differences in opioid and psychostimulant craving and relapse: a critical review

**DOI:** 10.1101/2021.03.30.21254644

**Authors:** Céline Nicolas, Natalie E. Zlebnik, Mehdi Farokhnia, Lorenzo Leggio, Satoshi Ikemoto, Yavin Shaham

## Abstract

A widely held dogma in the preclinical addiction field is that females are more vulnerable than males to drug craving and relapse. Here, we first review clinical studies on sex differences in psychostimulant and opioid craving and relapse. Next, we review preclinical studies on sex differences in psychostimulant and opioid reinstatement of drug seeking after extinction of drug self-administration and incubation of drug craving (time-dependent increase in drug seeking during abstinence). We also discuss ovarian hormones’ role in relapse and craving in humans and animal models and speculate on brain mechanisms underlying their role in cocaine craving and relapse in rodent models. Finally, we discuss imaging studies on brain responses to cocaine cues and stress in men and women.

The results of the clinical studies reviewed do not appear to support the notion that women are more vulnerable to psychostimulant and opioid craving and relapse. However, this conclusion is tentative because most of the studies reviewed were correlational, not sufficiently powered, and/or not a priori designed to detect sex differences. Additionally, fMRI studies suggest sex differences in brain responses to cocaine cues and stress. The results of the preclinical studies reviewed provide evidence for sex differences in stress-induced reinstatement and incubation of cocaine craving, but not cue- or cocaine priming-induced reinstatement of cocaine seeking. These sex differences are modulated in part by ovarian hormones. In contrast, the available data do not support the notion of sex differences in craving and relapse/reinstatement for methamphetamine or heroin in rodent models.

## Introduction

Drug addiction is characterized by high relapse rates during abstinence ^1,2^. Over the last decades, investigators examined sex differences in human drug use and relapse (see **Box 1** for glossary of terms) ^3^. In the early 1990s, Kosten et al. ^4^ reported that women have more severe cocaine use problems, more cocaine use days, and shorter abstinence periods. In contrast, women had better outcomes at the 6-month follow-up ^4^. In the early 2000s, Elman et al. ^5^ reported that cue-induced cocaine craving is higher in women. In parallel, many preclinical studies since the 1990s reported that female rats are more sensitive than males to the rewarding effects of cocaine, assessed by drug self-administration and conditioned place preference models ^6–10^. For example, female rats acquire cocaine self-administration faster than male rats ^11^. Additionally, investigators reported sex differences in relapse to cocaine seeking, as assessed by extinction-reinstatement and incubation of drug craving models ^12–14^ (see below). These findings have led to the widely held dogma, especially in the preclinical addiction field, that across drug classes, females are more vulnerable to initiation and escalation of drug use and to relapse to drug use during abstinence ^12,13,15,16^.

The goal of our review is to critically examine evidence for sex differences in psychostimulant and opioid craving and relapse in humans, and in reinstatement of drug seeking and incubation of drug craving in rat models. We refer readers to excellent comprehensive reviews of the preclinical literature on sex differences in initiation and escalation of drug self-administration, and drug reward and withdrawal ^10,16–18^.

We first review clinical studies on sex differences in psychostimulant (cocaine, methamphetamine) and opioid (heroin) craving and relapse. Next, we review preclinical studies on sex differences in psychostimulant and opioid (heroin, fentanyl, oxycodone) reinstatement of drug seeking and incubation of drug craving. We also discuss the role of ovarian hormones in cocaine craving and relapse/reinstatement in humans and animal models. Next, we propose a mechanistic model of the role of ovarian hormones in mediating sex differences in cocaine relapse in rat models. Finally, we summarize results from several human imaging studies on the brain response to drug cues and stress in men and women. In **Tables S1-5** and **Figures 1–2**, we provide a summary of the studies reviewed, and in **Box 1**, we provide a glossary of terms (blue font in the text). In the supplementary online section, we describe the methods of the systematic review of the clinical and preclinical literature.

**Figure 1.**
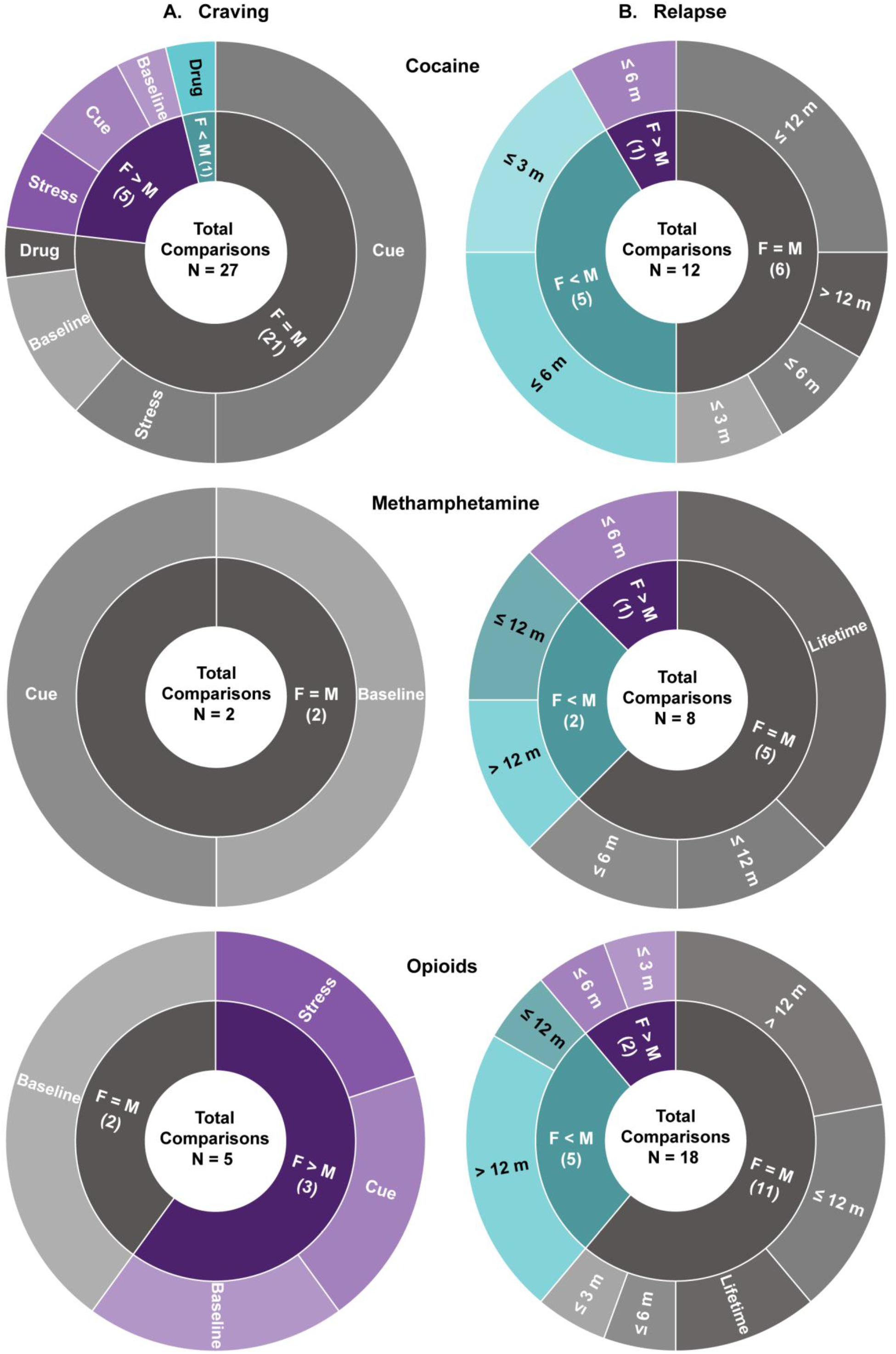
Clinical studies.

### Sex differences in drug craving and relapse: Clinical studies

We review studies on sex differences in psychostimulant and opioid craving and relapse in human laboratory and treatment settings (**Table S1, Figure 1**). Drug craving is correlated, to some degree, with drug use ^19–23^ and often predictive of future relapse ^22,24,25^. In human laboratory studies, drug craving is measured by psychometric self-report assessments before and after exposure to the drug itself, drug-related cues, or stressors ^25,26^. Cue-induced craving is achieved by exposing study participants to videos or pictures showing drug-associated cues (e.g., syringe), handling of drug paraphernalia, standardized scripts, or individualized guided imagery ^27^. Stress-induced craving is achieved by exposing study participants to psychological (e.g., personalized imagery, public speaking, mental arithmetic), physical (e.g., cold pressor), or pharmacological (e.g., alpha-2 adrenoreceptor antagonist yohimbine stressors ^25,28–30^.

In treatment settings, relapse is defined as resumption of regular drug use during outpatient treatment or after completion of inpatient or outpatient treatment. These studies rely on follow-up interviews at specified time points or ecological momentary assessment, real-time reporting of drug craving and use in the natural environment ^31,32^. We also review cross-sectional studies because they provide additional insight on sex differences in relapse vulnerability.

### Psychostimulants

#### Craving

##### Cocaine

Several studies examined sex differences in cocaine craving (**Table S1, Figure 1**), and while some studies suggest greater craving in women, other studies do not. Elman et al. ^33^ reported that when craving is assessed during early abstinence (12 h), women reported higher cocaine craving. Moran-Santa Maria et al. ^34^ reported that within 2-3 days of initiating abstinence, women report greater yohimbine-induced craving. Waldrop et al. ^35^ reported stronger correlation between peak craving and peak response to a social stressor (Trier Social Stress Test). In contrast, Brady et al. ^36^ reported that after similar abstinence length, men and women show similar craving when challenged with the same social stress test. Similarly, Back et al. ^37^ reported no sex differences in craving induced by psychological (Mental Arithmetic Task) or physical (Cold Pressor Task) stressors. Other studies found no sex differences in spontaneous (non-provoked) cocaine craving or cue-induced cocaine craving, two days into abstinence ^35^ or in non-abstaining users ^38^. Additionally, during early abstinence (< 7 days), craving induced by cocaine injection was either similar in men and women ^5^ or higher in men ^39^.

In contrast to some reports of sex differences in cocaine craving during early abstinence, when craving is monitored at least a week into abstinence, many studies reported no sex differences in spontaneous craving ^23,40,41^, cue-induced craving ^40,42^, or stress-induced craving ^43^. Additionally, there were no sex differences in cocaine craving in individuals with comorbid drug use disorder undergoing long-term opioid agonist treatment ^44,45^. However, Fox et al. ^46^ and Robbins et al. ^47^ reported higher cue-induced cocaine craving in women after 14-21 days of inpatient treatment or long-term outpatient treatment, respectively.

##### Methamphetamine

Tolliver et al. ^48^ reported no sex differences in cue-induced methamphetamine craving during early abstinence. Galloway et al. ^49^ reported no sex differences in spontaneous methamphetamine craving during 4 months of abstinence.

### Relapse

#### Cocaine

Kosten et al. ^4^ reported that upon entering treatment, women had higher relapse rates. However, they reported that there were no sex differences during the treatment study (desipramine or lithium carbonate), and at 6-month follow-up relapse rates were lower in women ^4^. In contrast, Kennedy et al. ^45^ reported that in polydrug (cocaine+heroin) users, women undergoing opioid agonist therapy showed higher relapse over a 7-month period. However, during 3-month ^50,51^ and 6-month ^52^ treatment follow-up, women showed lower relapse rates. Similarly, when relapse was assessed at treatment follow-up, women often exhibit better outcomes. Specifically, women had lower relapse rates at 6-month follow-up from both inpatient ^53^ and outpatient ^4^ treatment. In contrast, no sex differences in relapse were observed 9 months ^54^ or 12 months ^55,56^ after outpatient treatment. Additionally, during a 2-year period following a randomized clinical trial in polydrug (cocaine+opioid) users, men and women showed similar relapse rates ^57^.

#### Methamphetamine

Hillhouse et al. ^58^ reported higher relapse rates in women during 4-month outpatient treatment but no sex differences in relapse at 6-month or 12-month post-treatment. In contrast, Lanyon et al. ^59^ reported higher relapse risk in men at both 12-month and 5-year follow-ups. He et al. ^60^ and Brecht et al. ^61,62^ reported no sex differences in lifetime relapse rates in inpatients or after treatment completion, respectively.

### Opioids

#### Craving

Back et al. ^63^ reported greater spontaneous opioid craving in women and Yu et al. ^64^ reported higher cue-induced heroin craving in women during inpatient treatment. Additionally, Moran et al. ^65^ reported that over 4 months of outpatient opioid maintenance therapy, women showed higher stress-induced opioid craving. However, Herbeck et al. ^66^ reported no sex differences in opioid craving during 3 weeks of extended-release naltrexone treatment. Similarly, Kennedy et al. ^45^ reported no sex differences in heroin craving during 6 months of outpatient opioid agonist treatment.

#### Relapse

Maehira et al. ^67^ reported higher relapse rates in women during the first 2 months of abstinence. Ignjatova and Raleva ^68^ reported more heroin lapses in women during 6 months of opioid agonist treatment. In contrast, Kamal et al. ^69^ and Kennedy et al. ^45^ reported no sex differences in heroin relapse during the first 3 and 6 months of opioid agonist treatment, respectively.

After more prolonged abstinence periods (>1 year), there is some evidence for lower relapse in women. Gordon et al. ^70^ reported lower relapse rates for women at 1-year follow-up after treatment. Zimmer-Hofler ^71^ reported similar findings after 2 years of abstinence. Additionally, during a 2-year follow-up period during opioid agonist treatment for polydrug (cocaine+opioids) use, women had more drug-free days ^57^. Within the same longitudinal study, Darke et al. ^72,73^ reported that women were less likely to relapse over 3-year ^73^ and 11-year ^72^ follow-up periods. These results agree with previous studies reporting no sex differences in relapse at 7-year ^71^ and 8-year ^74^ follow-ups. Additionally, several studies reported no sex differences in relapse over 12-month follow-up during opioid agonist treatment ^75–77^, and no sex differences in lifetime opioid relapse rates ^78,79^.

## Conclusions

The studies reviewed do not support the notion of sex differences in drug craving and relapse for either psychostimulants or opioids. However, these studies suggest areas for further examination, including potential sex differences in craving and relapse vulnerability during early abstinence, where women may be more vulnerable.

### Sex differences in drug craving and relapse: Preclinical studies

Sex differences in drug relapse were examined using the reinstatement and incubation of drug craving models (**Table S2, Figure 2**) ^80^. In studies using the reinstatement model, investigators determined sex differences in drug priming-, drug cue-, and stress-induced reinstatement. In stress-induced reinstatement studies, the typical stressors investigators used in male rats were intermittent footshock ^81–83^ and yohimbine as a pharmacological stressor ^83,84^. In opioid users, yohimbine has been shown to induce stress- and withdrawal-like symptoms and opioid craving ^29^. However, interpretation of results from studies using yohimbine in reference to stress-induced reinstatement has been challenged by Chen et al. ^85^. They showed that at the dose range yohimbine is used in reinstatement studies, the drug’s effect on reinstatement is independent of the history of contingent self-administration and unrelated to the commonly assumed stress-like effects of yohimbine.

**Figure 2.**
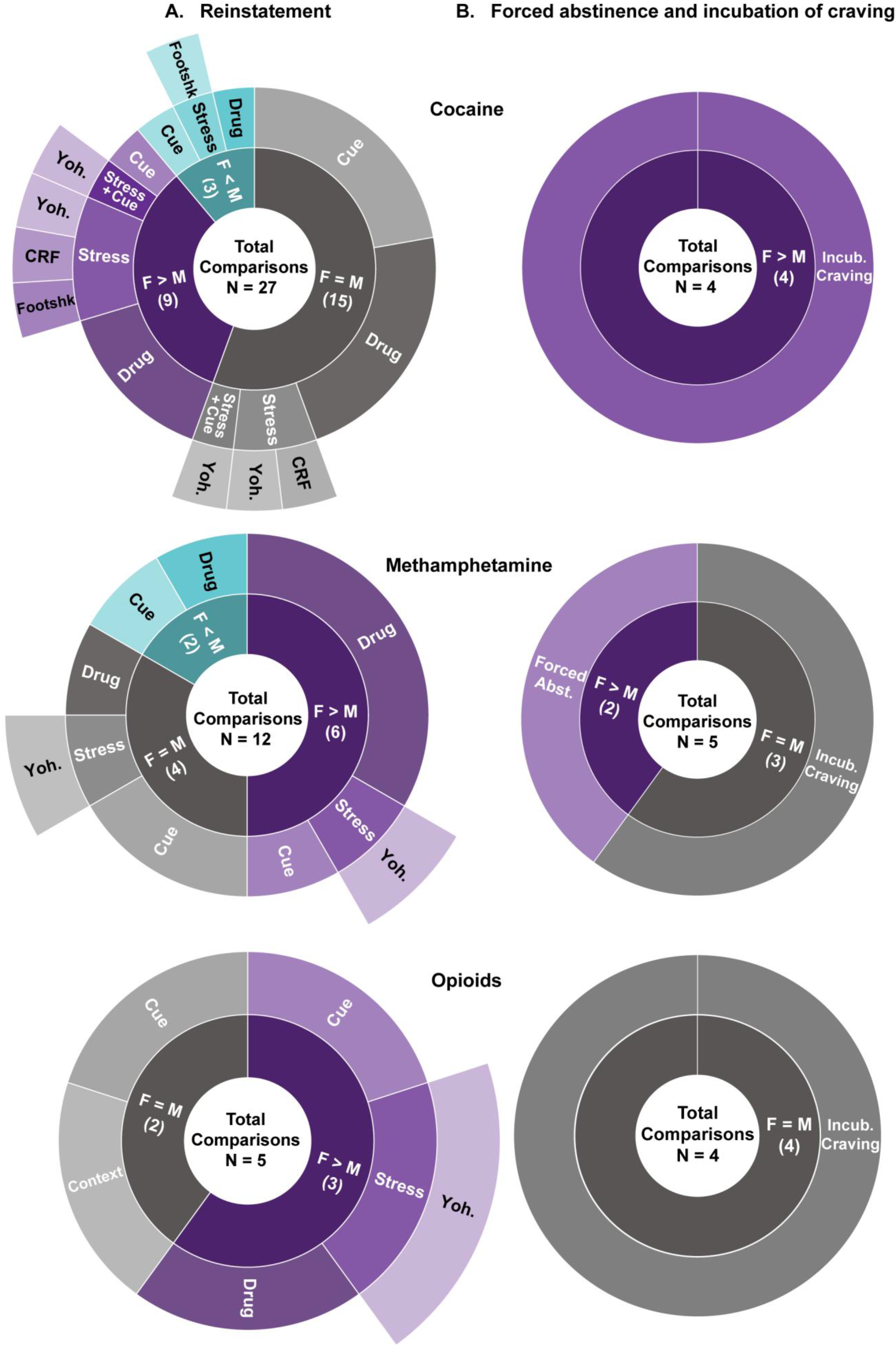
Preclinical studies.

### Psychostimulants

#### Drug priming-induced reinstatement

##### Cocaine

Lynch and Carroll ^86^ reported that female rats showed higher drug priming-induced reinstatement after limited-access continuous self-administration of cocaine at 1.0 and 3.2 mg/kg but not 0.32 mg/kg. This initial finding of sex differences in cocaine priming-induced reinstatement was confirmed in subsequent studies ^87–89^ (but see ref. Swalve et al. ^90^ for different results). In contrast, after extended-access continuous self-administration of cocaine, Zlebnik et al. ^91^ reported no sex differences in cocaine priming-induced reinstatement. Additionally, after extended-access intermittent self-administration, Kawa et al. ^92^ reported no sex differences in reinstatement induced by lower doses of cocaine priming (< 1.6 mg/kg, i.v.). A tentative conclusion from these studies is that females are more sensitive to cocaine priming-induced reinstatement after limited-access but not extended-access cocaine self-administration. Jordan and Anderson ^93^ reported that after limited access self-administration training with a high (0.75 mg/kg) but not low (0.25 mg/kg) cocaine unit dose that started on P28 for 30 days, non-reinforced responding during a single relapse test session after 30 abstinence days was higher in males after a priming cocaine injection (10 mg/kg, i.p). The relevance of these results to sex differences in cocaine priming-induced reinstatement after extinction is unknown because the rats did not undergo extinction training and the authors did not assess the effect of vehicle (saline) priming injections on drug seeking.

##### Methamphetamine

After both limited and extended access drug self-administration, methamphetamine (1 mg/kg) priming-induced reinstatement was higher in female rats ^94–97^. However, using the same priming dose, Everett et al. reported either no sex differences in methamphetamine-induced reinstatement ^98^ or higher reinstatement in males ^99^.

### Stress-induced reinstatement

#### Cocaine

Anker et al. ^100^ reported higher yohimbine-induced reinstatement and Feltenstein et al. ^101^ reported higher yohimbine+cue-induced reinstatement in females. In contrast, Zlebnik et al. ^91^ reported no sex differences in either yohimbine-induced or yohimbine+cue-induced reinstatement. Doncheck et al. ^89^ reported no sex differences in restraint stress-induced potentiation of drug priming-induced reinstatement. However, they reported that intermittent footshock potentiates drug priming-induced reinstatement in males but not females. In contrast, Connely et al. ^102^ reported higher footshock-induced reinstatement in females. Finally, in male rats, the effect of intermittent footshock stress on reinstatement of drug seeking is inhibited by extrahypothalamic corticotropin-releasing factor (CRF), and ventricular CRF injections mimic the effect of intermittent footshock on reinstatement ^82,83,103^. Based on these results, Buffalari et al. ^104^ examined sex differences in CRF-induced reinstatement of cocaine seeking. The main finding was that while ventricular injections of CRF reinstated cocaine seeking in both sexes, the response to CRF was more variable in females than in males.

#### Methamphetamine

Cox et al. ^95^ reported higher yohimbine-induced reinstatement in females, while Everett et al. ^98^ reported no sex differences ^98^. The reasons for these different results are unclear but may be due to a long abstinence period (21 d) before the extinction phase in the Everett study. As discussed above, interpretation of data from yohimbine studies in reference to stress-induced reinstatement is problematic ^83,85^.

### Cue-induced reinstatement

#### Cocaine

Zhou et al. ^105^ reported higher cue-induced reinstatement in females after limited-access continous cocaine self-administration. In contrast, under similar training conditions, several studies reported no sex differences in cue-induced reinstatement ^90,106–108^. Similarly, no sex differences in cue-induced reinstatement were observed after extended continuous ^91^ or intermittent access ^92^ cocaine self-administration.

### Incubation of craving and relapse after abstinence

#### Cocaine

Kerstetter et al. ^109^ reported longer lasting and higher incubation of cocaine craving (up to 180 days) in females after extended access continuous drug self-administration. We extended these results and reported higher incubation of cocaine craving in females after both continuous and intermittent extended access cocaine self-administration ^110^ (**Figure 3**). Johnson et al. ^111^ reported similar results with higher cocaine seeking in female rats tested at 1 or 30 abstinence days after continuous limited access cocaine self-administration. However, these results should be interpreted with caution because in both sexes, the incubation effect was variable and statistically non-significant, likely due to the use of the limited access procedure (incubation is less robust with this procedure ^112^). Madangopal ^113^ reported that incubation of the response to cocaine discriminative cues is more persistent in female rats and last for up to 200 abstinence days. However, these results should also be interpreted with caution because the study was not powered to detect sex differences.

**Figure 3.**
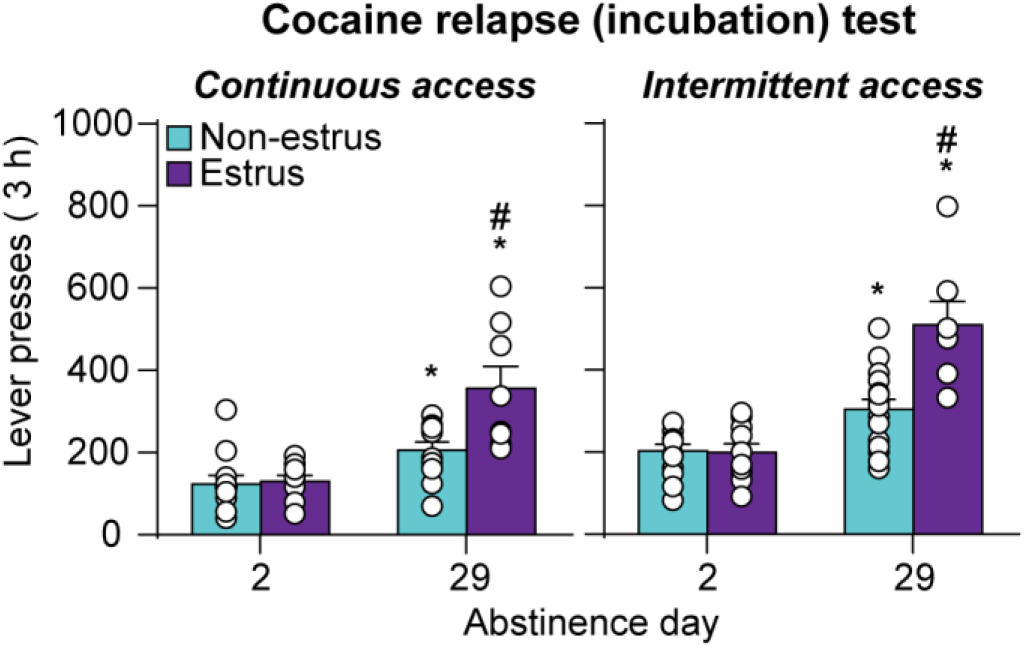
Role of estrous cycle in incubation of cocaine craving.

**Figure 4.**
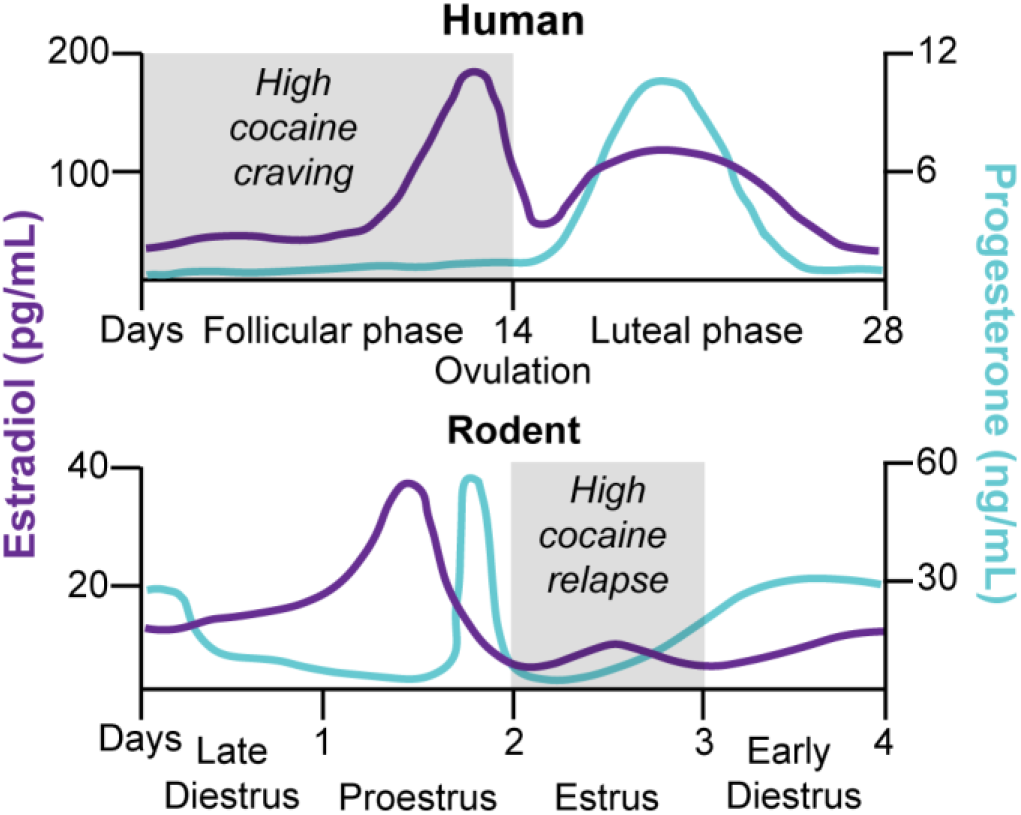
The female reproductive cycle in humans and rodents.

#### Methamphetamine

Venniro et al. ^114^ and Everett et al. ^98^ reported no sex differences in incubation of methamphetamine craving after either forced abstinence in homecage or voluntary abstinence in the drug self-administration chambers, the latter being achieved by providing rats with mutually exclusive choices between the self-administered drug and an alternative non-drug reward (palatable food or social interaction) ^115–119^. Similarly, Daiwile et al. ^120^ reported no sex differences in incubation of methamphetamine craving after forced abstinence. In contrast, Ruda-Kucerova et al. ^121,122^ reported that in a single relapse test after 15 days of forced abstinence from limited access methamphetamine self-administration, drug seeking was higher in female rats. In the context of sex differences in incubation of drug craving, these results are difficult to interpret because the authors did not establish that incubation had occurred under their experimental conditions.

### Opioids

#### Reinstatement after extinction

In an early study, Klein et al. ^123^ reported that reacquisition of oral fentanyl self-administration after extinction is higher in female rats. However, these results are difficult to interpret without ruling out potential confounds related to the oral route of drug administration (i.e., sex differences in pharmacokinetics and taste sensitivity). Smethells et al. ^124^ reported no sex differences in extinction responding but higher heroin priming- and yohimbine-induced reinstatement in females; they also reported higher heroin+cue- and yohimbine+cue-induced reinstatement in females. Vazquez et al. ^125^ reported higher extinction responding and cue-induced reinstatement of heroin seeking in females after limited access continuous drug self-administration in food-restricted rats. In contrast, in multiple replications, Bossert et al. ^126^ reported no sex differences in extinction responding, context-induced reinstatement, and reacquisition of oxycodone self-administration after extended access drug self-administration training. Bakhti-Suroosh et al. ^127^ tested both sexes for cue-induced reinstatement after 6 h of extinction in a single daily session performed 14 days after extended access intermittent fentanyl self-administration under two training conditions: FR1 schedule with or without 1-s timeout.

There were no sex differences in either extinction responding or cue-induced reinstatement in the no-timeout training condition. In contrast, sex differences (higher responding in females) emerged in the 1-s timeout training condition for extinction responding but not cue-induced reinstatement. The reasons for the sex-specific effect of the timeout manipulation on extinction responding are unknown.

#### Incubation of craving and relapse during abstinence

Unlike the mixed evidence for reinstatement of opioid seeking, there is no evidence for sex differences in incubation of opioid craving after either forced or voluntary abstinence, achieved via either providing the rats with alternative non-drug reward in a choice procedure or by introducing an electric barrier of increasing shock intensity near the drug-paired lever ^128^. Venniro et al. ^114^ reported no sex differences in either incubation of heroin craving after forced abstinence or reversal of incubation of heroin craving after food choice-induced abstinence. Venniro et al. ^129^ replicated the findings for forced abstinence and also reported no sex differences in the decrease in incubation of heroin craving after social choice-induced abstinence. Reiner et al. ^130^ reported no sex differences in relapse to fentanyl seeking after 2 weeks of food choice-induced abstinence. Fredriksson et al. ^131^ reported no sex differences in incubation of oxycodone craving after forced abstinence or potentiation of incubation of oxycodone craving after electric barrier-induced abstinence.

## Conclusions

The studies reviewed demonstrate sex differences for stress-induced reinstatement of cocaine seeking and incubation of cocaine craving after forced abstinence. In contrast, there is mixed evidence for sex differences in cue- or drug priming-induced reinstatement of cocaine seeking. In the case of cocaine priming, it appears that females are more vulnerable after limited-access but not extended-access cocaine self-administration. It is unlikely that these sex differences are due to pharmacokinetics, because there is no evidence for sex differences in brain and plasma levels of cocaine at the dose range used in reinstatement studies ^132^. Similarly, the evidence supporting sex differences in reinstatement of methamphetamine and opioid seeking across different reinstating stimuli is mixed at best. Additionally, there is consistent evidence for lack of sex differences in incubation of methamphetamine and opioid craving after forced or voluntary abstinence.

However, these conclusions should be confirmed in future studies, considering the relatively small number of sex differences studies, particularly for stress-induced reinstatement, where investigators primarily used yohimbine as the stress manipulation. In this regard, the subjects in the first intermittent footshock stress-induced reinstatement study were mostly female rats ^81^, but with few exceptions ^89,102,104,133^, only males were used in subsequent studies on reinstatement induced by intermittent footshock or other stressors like restraint, food restriction, or forced swim ^83,134^.

Finally, studies on reinstatement of food seeking also provide little evidence for sex differences. No sex differences were observed for sucrose priming-, yohimbine- and cue-induced reinstatement ^95,135,136^. However, Cox et al. ^95^ reported higher cue-induced sucrose reinstatement in female rats.

### Role of menstrual and estrous cycle

Several studies examined the role of ovarian hormones and menstrual/estrous cycle (**Figures 3–4**) in drug (primarily cocaine) craving and relapse in humans and rat models (**Tables S3-4**).

#### Clinical studies

Sofuoglu et al. ^137^ reported lower cocaine desire (craving) during the luteal than the follicular phase. Evans et al. ^138^ reported lower cocaine-induced subjective positive drug effects and craving during the luteal than the follicular phase. These results suggest a protective effect of progesterone (high during luteal phase) on cocaine-induced cocaine craving. In agreement with this idea, several studies reported that women with high endogenous progesterone levels (comparable to luteal phase) are less sensitive to stress- and cue-induced cocaine craving than women with low progesterone levels ^139–141^. Additionally, several studies reported that exogenous progesterone reduces positive subjective cocaine effects and craving in both women and men ^41,142–145^, confirming progesterone’s protective effects.

In contrast, in occasional intranasal cocaine users, Lukas et al. ^146^ reported no effect of the menstrual cycle on cocaine-induced subjective positive drug effects. The results of this negative study agree with those from other studies in cocaine-dependent women ^42,147^. Additionally, Fox et al. ^148^ reported no variations in craving over the menstrual cycle during the first month of abstinence from smoked cocaine, but the ratio of estradiol/progesterone was stable across the cycle, which could explain these negative results.

Both the negative and positive results reviewed above should be interpreted with caution because of relatively small sample sizes and the presence of other factors in these samples that may also play a role in the behavioral outcomes under investigation (e.g., severity of the cocaine use disorder, route of administration).

#### Preclinical studies

The results of studies on the role of estrous cycle and ovarian hormones in reinstatement and incubation of craving are summarized in **Table S4**.

##### Cocaine

Cocaine priming-induced reinstatement is higher during estrus than diestrus or proestrus ^109,149–151^. The expression of incubation of cocaine craving is higher during estrus than non-estrus (**Figure 3**) ^109,110^. In contrast, evidence for the role of estrous cycle in cue- and stress-induced reinstatement is mixed. Fuchs et al. ^107^ reported lower cue-induced reinstatement of cocaine seeking in females in estrus than non-estrus. Feltenstein et al. ^101^ reported that yohimbine+cue-induced reinstatement is lower in estrus and diestrus than proestrus. In contrast, they reported that the estrus phase has no effect on yohimbine-induced reinstatement of cocaine seeking. Peterson et al. ^152^ and Bechard et al. ^108^ also reported similar cue-induced reinstatement of cocaine seeking during different phases of the estrous cycle.

Suppression of ovarian hormones by ovariectomy decreases cocaine priming-induced reinstatement, while chronic estradiol treatment in ovariectomized rats restores this reinstatement to levels similar to those of sham rats ^153,154^. Additionally, acute proestrus-level estradiol in ovariectomized rats potentiates cocaine priming-induced reinstatement ^155^. In contrast, exogenous progesterone treatment in free-cycling rats decreases cocaine priming-induced reinstatement ^149,153^. Anker et al. ^153^ also reported that exogenous progesterone treatment in ovariectomized rats inhibits the facilitating effect of estradiol on cocaine priming-induced reinstatement. Together, these results suggest a role of ovarian hormones in cocaine relapse with estradiol increasing relapse vulnerability, while progesterone having an opposite effect.

##### Methamphetamine and heroin

The role of ovarian hormones in relapse/reinstatement to methamphetamine and heroin seeking is largely unknown. Cox et al. ^95^ reported that the estrous cycle has no effect on methamphetamine priming-induced reinstatement. Vazquez et al. ^125^ reported that estradiol or progesterone treatment has no effect on cue-induced reinstatement of heroin seeking. Sedki et al. ^133^ reported that food restriction for 2 weeks (a chronic stressor) increases relapse to heroin seeking after homecage forced abstinence in female rats. This effect is not decreased by ovariectomy; unexpectedly, in ovariectomized rats, estradiol replacement but not progesterone injections decrease the potentiation effect of chronic food restriction on relapse. In contrast, Bakhti-Suroosh et al. ^127^ reported higher extinction responding and cue-induced reinstatement of drug seeking after 14 days of forced abstinence from extended intermittent access fentanyl self-administration during estrus vs. non-estrus.

## Conclusions

The clinical and preclinical studies reviewed above suggest that under certain conditions, cocaine craving and relapse are dependent on the menstrual/estrous cycle, with higher vulnerability during the follicular/estrus phase. The hypothesis that emerges from these studies is that progesterone decreases cocaine craving and relapse vulnerability, while estradiol has opposite effects. The results of Sedki et al. ^133^ described above on the ‘protective’ effect of estradiol suggests that this hypothesis may not generalize to opioid drugs (but see Bakhti-Suroosh et al. ^127^ for results congruent with the notion of opposing roles of estradiol and progesterone in drug relapse).

### Brain mechanisms

The brain mechanisms involved in the putative opposite effects of estradiol and progesterone on cocaine craving and relapse are unknown. We speculate that the mesolimbic dopamine system, with its projections from ventral tegmental area (VTA) to nucleus accumbens (NAc), is critically involved. This system plays important roles in both reinstatement of cocaine seeking after extinction ^156–158^ and incubation of cocaine craving ^159,160^. Many studies reported that amphetamine-induced striatal and accumbal dopamine release is increased during estrus in free-cycling females ^161–163^ and by estradiol treatment in ovariectomized rats ^164,165,166, 167,168^. In contrast, progesterone has an opposite effect by inhibiting striatal dopamine release in estradiol-primed ovariectomized females ^165,169^. Calipari et al. ^170^ also reported that dopamine activity in the VTA-to-NAc projection is higher during estrus than non-estrus.

We propose that estradiol and progesterone exert opposite effects on dopamine neurotransmission in VTA-to-NAc projection, leading to increased cocaine craving and relapse by estradiol and decreased cocaine craving and relapse by progesterone (**Figure 5**). Other neurobiological mechanisms of reinstatement and incubation of cocaine craving, including mesocorticolimbic glutamate transmission ^159,160,171–173^, may also contribute to the sex differences described above. As discussed elsewhere, there are sex differences in brain glutamate systems and drug-induced neuroadaptations in glutamate transmission that may play a role in this regard ^174^.

**Figure 5.**
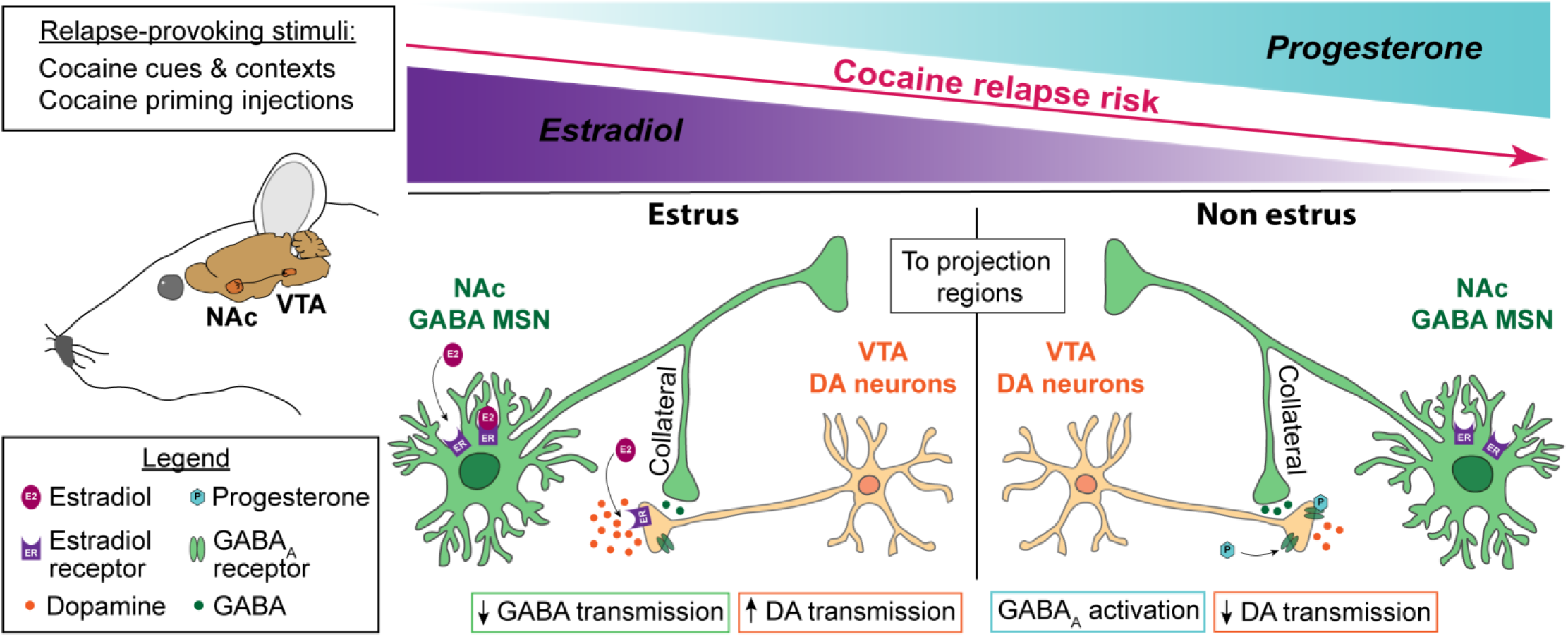
Proposed neuronal mechanism of the role of estrous cycle in cocaine relapse in rat models.

#### Human imaging studies on sex differences in response to cocaine cues and stress

During the last several decades, numerous studies have used positron emission tomography (PET) ^175^ and functional magnetic resonance imaging (fMRI) ^176^ to identify brain regions activated (or inhibited) during craving induced by drug priming ^177^, drug cues ^178^, and stress ^179^. However, with rare exceptions, these studies were either performed in male subjects or included both sexes but were not designed (lacked power) to assess sex differences. Indeed, in our systematic literature search, we only identified **six** studies that statistically evaluated sex differences in brain activity during cue- or stress-induced cocaine craving (**Table S5**). We describe these studies below. In the Supplemental section we also describe results from several studies where investigators separately analyzed imaging data within each sex. The data from these studies are inconclusive and difficult to interpret without knowing whether the interactions between sex and cue or stress conditions are significant (see Nieuwenhuis et al. ^180^).

In the studies described below, cue-induced craving was provoked by either presentation of videos or pictures showing cocaine-associated cues or by standardized or personalized scripts ^38,181,182^. Stress-induced craving was induced by standardized or personalized scripts ^28,43^. Brain activity changes induced by cocaine or stress cues were compared to those induced by neutral cues. Participants in these studies were either abstinent and underwent inpatient or outpatient treatment ^43,181,182^ or were non-treatment seeking active cocaine users ^38^.

Kilts et al. ^181^ used PET with [^15^O]H2O to measure regional cerebral blood flow (an index of brain activity) after cue exposure during early abstinence (days 1-14) in eight women. They compared the women to five men from their previous study ^183^ and three new men. There were no sex differences in cue-induced cocaine craving. Exposure to cocaine cues induced stronger activation (increased glucose utilization) in right amygdala, left insula, right postcentral gyrus, and left caudate nucleus in men. In contrast, women showed increased activation of right precentral gyrus, middle frontal gyrus, and posterior cingulate gyrus. These results should be interpreted with caution because of the low number of participants and the use of male data from a previous study.

Volkow et al. ^38^ used PET with 2-deoxy-2[18F]fluoro-D-glucose (^18^FDG) to measure brain glucose metabolism in response to cocaine cues in ten women and sixteen men. There were no sex differences in cue-induced cocaine craving. In contrast, sex differences were detected in cue-induced whole brain metabolism (glucose utilization) with significant decreased activity in women and modest increased activity in men. Additionally, analyses of specific brain areas showed decreased glucose utilization in frontal, cingulate and parietal cortices, thalamus, and midbrain in women. In contrast, men showed increased glucose utilization in right inferior frontal gyrus. Direct comparisons of men and women showed sex × cue (cocaine cue, neutral cue) interaction due to greater decrease in glucose utilization among women in frontal (broca areas 8, 9, 10), anterior cingulate, posterior cingulate, inferior parietal, and dorsomedial thalamus.

The reasons for the different pattern of results in the two aforementioned studies, cue-induced brain activation in women in Kilts et al. ^181^ vs. cue-induced inhibition in Volkow et al. ^38^ are unknown. One potential reason is that subjects in the first study ^181^ were treatment seekers tested during abstinence, while those in the second study ^38^ were non-treatment seeking tested during active cocaine use.

Kober et al. ^184^ investigated fMRI-based changes in brain activity in response to videos depicting cocaine use, gambling, or sad scenarios in participants with cocaine dependence or pathological gambling. There were no sex differences in cocaine craving or urges between cocaine-dependent (n=18) men and women (n=12). However, men showed greater dorsomedial prefrontal cortex and superior frontal gyrus activation in response to the cocaine videos.

Joseph et al. ^185^ used fMRI to investigate the effects of oxytocin on cocaine craving and cocaine cue-induced activity in right amygdala and dorsomedial PFC among cocaine-dependent participants with (24 men, 16 women) or without (19 men, 8 women) childhood trauma history. Independent of the trauma condition, there were no sex differences in cue-induced cocaine craving, and in both sexes, oxytocin had no effect on this measure. Independent of the trauma condition, there were no sex differences in the placebo groups for cue-induced activation of dorsomedial PFC, a finding different from that of Kober et al. ^184^, and oxytocin decreased this activation in all groups in a sex independent manner. A different pattern of results was observed for cue-induced activation of right amygdala, where sex differences (activation in men but not women) were observed in the trauma but not non-trauma participants. Additionally, in the trauma groups, oxytocin increased cue-induced right amygdala activity in women but decreased this activity in men. In contrast, oxytocin had no effect on cue-induced right amygdala activity in men and women without trauma. The small sample size of women with prior trauma limits the interpretation of the sex-specific effects of oxytocin on amygdala activity in this study.

Zhang et al. ^182^ used fMRI to investigate periaqueductal gray (PAG) activity and connectivity between PAG and ventromedial prefrontal cortex (vmPFC) after craving induced by cue exposure during early abstinence (7-10 days) in ten women and forty-two men. The PAG is best known for its role in pain, avoidance, and defensive behaviors ^186,187^. There were no sex differences in cue-induced cocaine craving, cue-induced PAG activity, or cue-induced increased PAG-vmPFC connectivity.

However, PAG-vmPFC connectivity strength was positively correlated with cue-induced craving in men but negatively correlated in women. These results suggest sex differences in the role of PAG-vmPFC connectivity in cue-induced cocaine craving. However, the study’s results should be interpreted with caution because of the low number of women subjects and other significant sex differences in the sample (age and depression score).

Li et al. ^43^ investigated brain activation using fMRI during stress imagery in ten women and seventeen males who were abstinent for 2-3 weeks. There were no sex differences in stress-induced cocaine craving. Additionally, there were no sex differences in the observed negative correlations between stress-induced craving and activation of the anterior and posterior cingulate. However, sex differences (higher activation in women) were observed for stress-induced activation of left frontolimbic areas, including anterior cingulate, insula, dorsolateral and medial PFC, inferior frontal cortices, and posterior cingulate cortex.

## Conclusions

The results of the studies reviewed demonstrate strong sex differences in the effect of cue- and stress-induced cocaine craving manipulations on brain activity, with both quantitative (different degrees of activation) and, unexpectedly, qualitative (opposite effects) differences (**Figure 6**). These differential brain responses occurred despite the consistent lack of sex differences in cue- or stress-induced subjective craving. Thus, a tentative conclusion from these studies is that to the degree that correlational results from imaging studies reflect causes rather than consequences of drug craving, the brain circuits controlling cocaine craving in men and women are likely different. The studies reviewed also suggest that in both sexes the brain mechanisms of cue-induced vs. stress-induced cocaine craving are largely dissociable. This conclusion is consistent with results from preclinical studies on differences in brain circuits of cue- vs. stress-induced reinstatement of drug seeking ^134,156,188^. Another observation from the studies reviewed is that cue-induced brain activation vs. inhibition is dependent on the addiction phase (active cocaine use vs. abstinence) ^38,181^.

**Figure 6.**
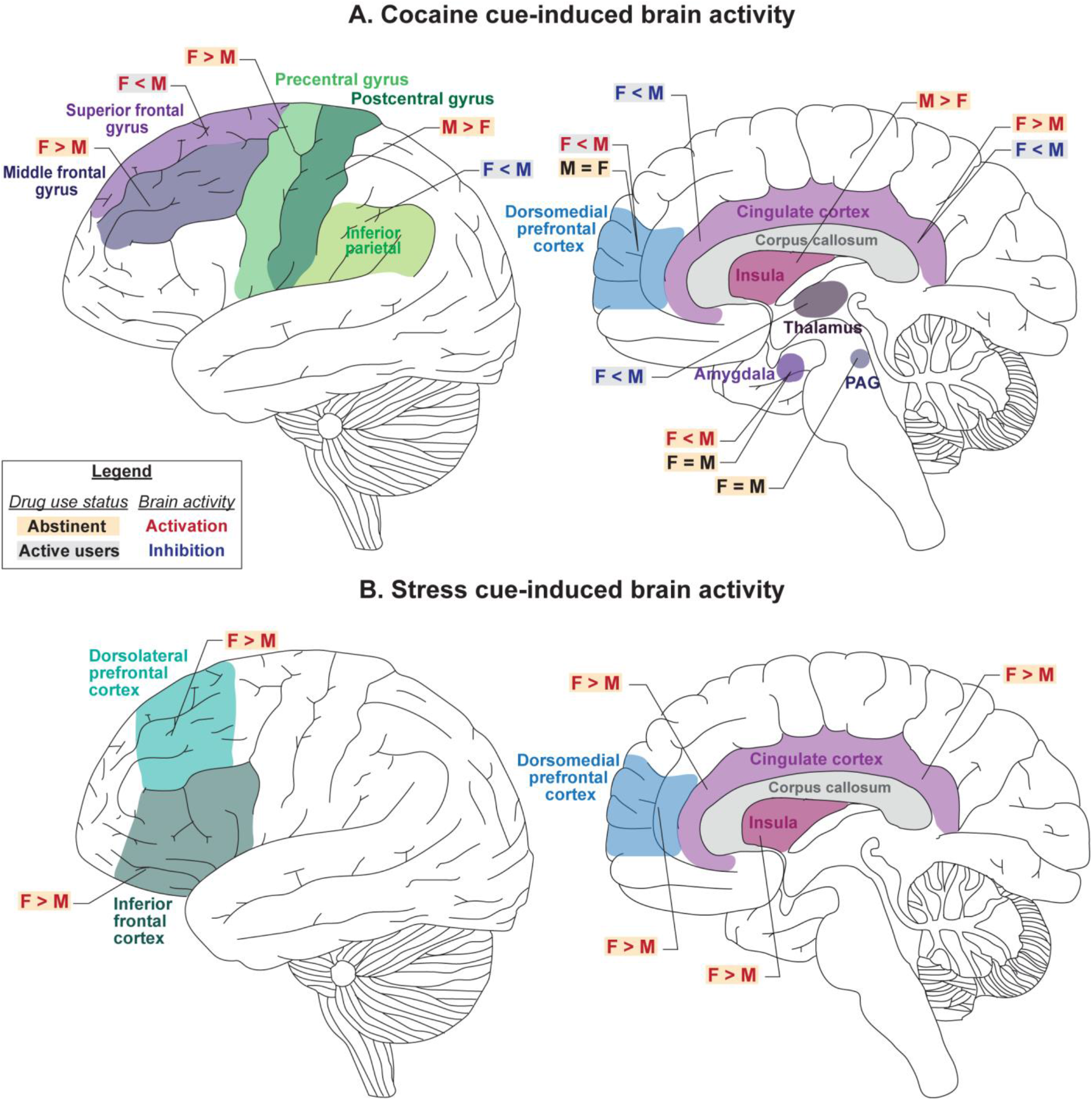
Sex differences in cocaine cue- and stress-induced brain activation.

A limitation of the studies reviewed, particularly in reference to an important clinical outcome— relapse after prolonged abstinence—is that they were performed during early abstinence or ongoing cocaine use. Thus, the pattern of brain activation in both sexes in response to drug cues and stress during protracted abstinence is unknown. Finally, for the fMRI studies, a common scheme in the studies reviewed was small sample sizes that are well below the recommended number of human subjects required for reproducible task-based fMRI studies ^189^. Thus, future research should replicate these studies with larger sample sizes.

### Conclusion and clinical implications

A prevailing notion in the preclinical addiction field is that females are more vulnerable than males to drug addiction and relapse across drug classes ^12,13,15^. Here, we critically reviewed this notion with respect to psychostimulant and opioid craving and relapse in humans and rat models. Unexpectedly, as our own research was guided by this notion ^91,110^, our review does not support ubiquitous female susceptibility to craving and relapse.

Our main conclusion of the clinical literature is that the published studies do not support the idea that women are more vulnerable to psychostimulant and opioid craving and relapse. However, this conclusion is tentative because many of the studies reviewed were either correlational or not sufficiently powered to detect sex differences.

Our main conclusion of the preclinical literature is that there are sex differences in stress-induced reinstatement and incubation of cocaine craving, which in the latter case are modulated, in part, by ovarian hormones. These data do not support the hypothesis that ovarian hormones play a critical role in the initiation of drug use and enhancement of the initial reinforcing effects of cocaine in females, but not the other phases of the addiction process ^14^. In contrast, there is minimal evidence for sex differences for either cue- or cocaine priming-induced reinstatement of cocaine seeking. Additionally, the studies with methamphetamine and heroin (and other opioid drugs) do not support the notion of sex differences in reinstatement of drug seeking or incubation of craving for these drugs. However, for reinstatement of methamphetamine and heroin seeking, our conclusion is tentative, because only a few studies were published, and the results are mixed.

The reasons for the drug-specific evidence of sex differences in rodent models are unknown. Potential reasons could be the distinct neurobiological mechanisms of relapse between opioids vs. psychostimulants and cocaine vs. methamphetamine ^188,190^, and the differential interaction of these drugs with ovarian hormones or organizational sex effects.

Another main conclusion is that fluctuations in ovarian hormones appear to play a role in cocaine craving in humans, as well as cocaine priming-induced reinstatement and incubation of craving in rat models. The hypothesis that emerges from these studies is that progesterone decreases drug craving and relapse, while estradiol has an opposite effect (see **Figure 5** for a proposed brain mechanism for these effects).

An issue to consider from a translational perspective is the apparent lack of concordance between preclinical and clinical studies with cocaine, with some evidence supporting sex differences in rat models, but not in humans. This discrepancy is not surprising, because of the complex social, legal, cultural, and language-related factors that contribute to human addiction that cannot be modelled in rodents ^191–193^. However, a close inspection of the human results suggests some similarities, opportunities for future research, and potential treatment implications. Specifically, in some studies women report greater cocaine craving than men during early abstinence ^33–35^. Additionally, several laboratory studies report decreased cocaine craving during the luteal vs. follicular phase ^137,138^. Thus, abstinence attempts during the luteal phase may be more successful than those during the follicular phase. Indeed, similar approaches have led to favorable cessation outcomes in tobacco smokers ^194–196^. Additionally, treatment with exogenous progesterone reduced cocaine craving ^41,142–145^. Together, these results suggest that in both humans and rodent models, the menstrual/estrous cycle contributes to cocaine seeking and highlight progesterone as a potential adjunct pharmacotherapy to reduce cocaine relapse.

The influence of gonadal hormones on drug craving is an example of how different mechanisms may promote susceptibility to relapse in males and females. Additionally, as discussed above, despite the apparent lack of sex differences in self-reported craving and rates of relapse in humans, neuroimaging studies identified sex differences in regional brain activation associated with cue- and stress-induced cocaine craving (**Figure 6**) ^38,42,43,181,182^. These results suggest sex differences in brain mechanisms of cocaine (and potentially other drugs) craving and potentially relapse. This conclusion is supported by preclinical studies suggesting sex-specific mechanisms of cocaine-seeking behaviors ^89,170^. Variation in the underlying neurobiology of craving and relapse between men and women may result in differential treatment effects. For example, women experienced a greater reduction in stress- and cue-mediated cocaine craving than men after administration of the alpha2 adrenergic agonist guanfacine ^46^. Sex differences in ‘treatment’ effects were also demonstrated for drug-seeking behavior in rodent models ^87,90,96,100,197,198^. For example, in a recent study Bossert et al. ^126^ reported that female rats are less sensitive than males to the effect of a mu opioid receptor partial agonist on context-induced reinstatement and reacquisition of oxycodone self-administration.

Finally, our review focused on degree and prevalence of self-reported craving and rates of relapse and found no clear evidence for sex differences. However, the etiology of these phenomena may differ between men and women ^63^, and future studies should examine the complex factors that may differentially promote craving and relapse in both sexes.

## Data Availability

N/A

## Figure legends

**Figure 1**. *Summary of clinical studies on sex differences in psychostimulant and opioid craving and relapse*. Both (**A**) craving and (**B**) relapse panels depict the proportion of observations and list the total number of comparisons of sex differences in which F > M, F = M, and F < M. Conditions under which craving was measured and the abstinence period when relapse was assessed is also displayed. <u>Abbreviations:</u> F = females, M = males, m = months. <u>Note:</u> the number of comparisons does not equal the number of studies for a given category in Table S1, because in some studies investigators assessed more than one dependent measure (e.g., both cue- and stress-induced craving).

**Figure 2**. *Summary of preclinical literature on sex differences in psychostimulant and opioid reinstatement and incubation of craving*. Both (**A**) reinstatement of drug-seeking behavior and (**B**) incubation of craving and drug seeking after forced abstinence panels depict the proportion of observations and list the total number of comparisons of sex differences in which F > M, F = M, and F < M. The conditions under which drug seeking was measured are also displayed. <u>Abbreviations:</u> F = females, M = males, Yoh = yohimbine, CRF = corticotropin-releasing factor. <u>Note:</u> the number of comparisons does not equal the number of studies for a given category in Table S2, because in some studies investigators assessed more than one dependent measure (e.g., both cue- and stress-induced reinstatement).

**Figure 3**. *Effect of estrous cycle on incubation of craving after continuous-access and intermittent-access cocaine self-administration in female rats*. Relapse (incubation) test. Mean ± SEM number of active lever presses per session after continuous (Non-estrus: n = 12 for day 2 and 29, estrus: n = 9/8 for day 2/29) and intermittent (Non-estrus: n = 13/16 for day 2/29, estrus n =10/7 for days 2/29) access drug self-administration. * Different from day 2 within each estrous phase, p < .05; # Different from non-estrus on day 29, p < .05. Adapted from Nicolas et al. ^110^.

**Figure 4**. *Schematic comparison of the menstrual cycle in humans and the estrous cycle in rodents and fluctuation of estradiol and progesterone levels across the cycle phases*. In humans, the menstrual cycle is divided into the follicular and luteal phases separated by ovulation ^199,200^. The cycle begins with menses, and the onset of the follicular phase is characterized by high levels of estradiol, with a peak during the preovulation period before decreasing to a moderate level during the luteal phase. Conversely, progesterone is at its lowest level during the follicular phase and starts increasing during the preovulation period to peak at the midluteal phase. In female rodents, the estrous cycle is divided into late diestrus, proestrus, estrus, and early diestrus phases with ovulation occurring between proestrus and estrus ^201,202^. Estradiol peaks twice at the middle of the early diestrus and proestrus phases and drops by ovulation. Progesterone peaks at the end of the early diestrus and proestrus phases, and low stable levels remain for the rest of the cycle. In many studies, investigators pool together early and late diestrus phases to a single phase called diestrus because of similar hormonal and cytological characteristics. Additionally, when no behavioral differences are observed during proestrus and diestrus, investigators combine these phases and call the combined phase nonestrus. There are major differences in the reproductive cycle of women and female rodents: the duration of the cycle, the cycle pattern of estradiol and progesterone, and the amplitude of hormone level variations. However, when the hormonal ratio over the phases is compared, some analogies are conventionally made: the follicular phase is comparable to the estrus phase (progesterone < estradiol) and the luteal phase to the nonestrus phase (progesterone > estradiol) ^200,202^.

**Figure 5**. *A proposed brain mechanism model of the role of estradiol and progesterone in cocaine relapse in rodent models*. There is evidence that estradiol potentiates striatal and nucleus accumbens (NAc) dopamine release by modulating GABAergic neurotransmission of medium spiny neurons (MSN) through collaterals synapsing on dopamine neurons ^202,203^. This effect is likely mediated by membrane estrogen receptors (membrane-associated ERα and membrane-associated ERβ) expressed in MSNs ^204^.

ERα and ERβ antagonists prevent estradiol enhancement of amphetamine-induced dopamine release ^205^ and overexpression of ERα in the striatum increases the effect of estradiol on K+ induced GABA release ^206^. Additionally, ERα and ERβ are expressed on VTA dopamine neurons terminals in NAc ^203^, which play a critical role in reinstatement of cocaine seeking ^157,207^. Together, these results suggest a role of ERα and ERβ in estradiol regulation of dopamine neurotransmission and, by implication, in reinstatement of cocaine seeking. Additionally, increased dopamine release by estradiol would be at least in part to due to its action on dopamine receptors: striatal dopamine receptor 2 (Drd2) affinity decreases during estrus ^208^, and estradiol injections in ovariectomized rats decrease Drd2 binding in striatum ^209^.

In contrast to estradiol, progesterone decreases striatal dopamine release, as shown in estradiol-primed ovariectomized females treated with progesterone ^165,169^. Progesterone and its metabolites are positive allosteric modulators of GABA_A_ receptors ^210–212^. Drugs that promote GABA_A_ function (e.g., imidazenil, diazepam) decrease cocaine-induced increases in dopamine release in NAc shell ^213^. Consequently, progesterone could protect against estradiol-induced increases in cocaine seeking by inhibiting NAc dopamine release via increased GABA_A_ receptor transmission.

Together, we propose that during the estrus/follicular phase, cocaine priming- or cocaine cue-induced NAc dopamine release is increased by estradiol through its action on ER in GABAergic medium spiny striatal neurons and/or dopamine neurons terminals, leading to disinhibition of dopamine neurons and/or directly enhanced VTA dopamine cell firing via decreased Drd2 signaling, resulting in increased cocaine reinstatement/relapse. In contrast, during non-estrus/luteal phase, high progesterone levels may inhibit dopamine release induced by drug priming or drug cues through its action on GABA_A_ receptors expressed in VTA dopamine terminals ^214,215^, resulting in decreased cocaine relapse.

**Figure 6**. *Sex differences in cue- and stress-induced brain activation in cocaine-dependent subjects*. Schematic illustration of sex differences in (**A**) cocaine cue-induced and (**B**) stress-induced brain activation (assessed by PET or fMRI). PAG: periaqueductal gray, M: male, F: female.

**Box 1. Glossary of terms.**

**Conditioned place preference:** A Pavlovian conditioning procedure in which one distinct context is paired with noncontingent drug injections while another context is paired with vehicle injections. During subsequent drug free tests, increased preference for the drug context serves as a measure of the drug’s rewarding effects.

**Continuous drug self-administration:** A drug self-administration procedure in which the drug is continuously available for the duration of the daily sessions. This procedure results in high and stable drug concentrations in the brain.

**Craving**: An affective state described as an urge for drug; it can be induced in human drug users by exposure to the self-administered drug, drug cues and context, or stress.

**Drug self-administration**: An operant procedure in which laboratory animals lever press (or nose poke) for drug injections or oral drug delivery. Most (but not all) drugs abused by humans are self-administered by rodents and nonhuman primates.

**Extinction**: The decrease in the frequency or intensity of learned responses after the removal of the unconditioned stimulus (e.g., drug) that has reinforced the learning. In studies on incubation of drug craving and relapse after forced or voluntary abstinence, extinction responding (in the presence of the drug-paired contextual and discrete cues) is the operational measure of drug seeking.

**Incubation of drug craving**: A hypothetical psychological process inferred from the findings of time-dependent increases in non-reinforced drug seeking after cessation of drug self-administration in rodents.

**Intermittent drug self-administration:** A drug self-administration procedure in which the drug is repeatedly available for short periods that are separated by long timeout periods (typically 6 to 12 cycles of 5 min drug access, 25 min timeout). Exposure to this procedure induces binge-like self-administration behavior and spiking brain drug levels.

**Reinstatement of drug seeking**: Post-extinction resumption of operant behavior that had previously been maintained by a drug. Reinstatement is induced by a priming drug injection, stressors, contexts previously paired with drug self-administration, or response-contingent presentation of drug-associated cues.

**Relapse**: Resumption of drug-taking behavior during self-imposed (voluntary) or forced abstinence in humans and laboratory animals.

**Sex**: Characterization of an individual as female or male from biological and morphological features.

## Supplemental online material

Methods of the systematic review

Figure S1: Flow chart of the literature search of the clinical studies Figure S2: Flow chart of the literature search of the preclinical studies Additional human imaging studies on sex differences in brain activity

Table S1. Psychostimulant and opioid craving and relapse: clinical studies

Table S2. Psychostimulant and opioid reinstatement and incubation of craving: preclinical studies Table S3. Role of menstrual cycle on cocaine subjective effects and craving

Table S4. Role of estrous cycle in cocaine reinstatement and incubation of craving

Table S5. Summary of human brain imaging studies

