## Supplementary Online Material for "Sex differences in opioid and psychostimulant craving and relapse: a critical review"

3-30-2021

medRxiv (systematic review)

Table of contents

Methods of the systematic review

Figure S1: Flow chart of the clinical literature search and review

Figure S2: Flow chart of the preclinical literature search and review

Additional human neuroimaging studies on sex differences

Table S1. Psychostimulant and opioid craving and relapse: clinical studies

Table S2. Psychostimulant and opioid reinstatement and incubation of craving: preclinical studies

Table S3. Role of menstrual cycle in cocaine subjective effects and craving

Table S4. Role of estrous cycle in cocaine reinstatement and incubation of craving

Table S5. Human neuroimaging studies on cocaine cue- and stress-induced brain activity

**Methods of the systematic review**

**Clinical studies**

The literature search strategy was developed and executed by Ms. Diane Cooper (an NIH informationist) in consultation with the authors. Consistent with the Preferred Reporting Items for Systematic Reviews and Meta-Analyses (PRISMA) guidelines, we searched MEDLINE, Embase, and PsycINFO with no time limits (see below). We included original research studies that evaluated sex/gender differences in psychostimulant and/or opioid craving and/or relapse in adults. We excluded preclinical studies, review articles, commentaries, case reports, and conference abstracts. We also excluded studies that reported the results in women and men without formal statistical comparisons. NEZ and MF independently screened the titles/abstracts and filtered the records according to the eligibility criteria described above. They compared the two resulting lists and discussed discrepancies. In case of disagreement, they consulted LL and reached a consensus. Following this initial screen, we retrieved the full texts of potentially eligible records. Using an identical process as the title/abstract stage, we evaluated the full text of these articles for final inclusion in this review (**Figure S1**). We used EndNote X9 to collect, de-duplicate, and manage the records. Below we provide the details of the search strategy for MEDLINE, which we also adapted for the other two databases (Embase and PsychINFO) using similar terms:

("craving"[MeSH] OR "craving"[tiab] OR "recurrence"[MeSH] OR "relapse"[tiab] OR "abstinence"[tiab] OR "remission"[tiab]) AND ("analgesics opioid"[Pharmacological Action] OR "Central Nervous System Stimulants"[Pharmacological Action] OR "Cocaine"[MeSH] OR "Amphetamine-Related Disorders"[MeSH] OR "Opioid-Related Disorders"[MeSH] OR "Cocaine-Related Disorders"[MeSH] OR "opioid"[tiab] OR "opiate"[tiab] OR "opium"[tiab] OR "heroin"[tiab] OR "morphine"[tiab] OR "stimulants"[tiab] OR "amphetamine"[tiab] OR "dextroamphetamine"[tiab] OR "methamphetamine"[tiab] OR "Cocaine"[tiab] OR "caffeine"[tiab]) AND ("sex characteristics"[MeSH] OR "sex factors"[MeSH] OR "sex characteristics"[tiab] OR "sex differences"[tiab] OR "gender"[tiab] OR “gender differences”[tiab]). Filters: Humans

**Preclinical studies**

The preclinical literature search was consistent with the clinical search and based on methods Shaham and colleagues used in previous reviews of the preclinical literature of studies using animal models of relapse and craving (e.g., ref. ^1-6^). In accordance with the PRISMA guidelines, Ms. Diane Cooper searched MEDLINE, Embase, and PsycINFO, using keywords and controlled vocabulary terms relevant to the concept of the review, with no time limits. Below we provide the details of the research strategy for MEDLINE, which we also adapted for the other two databases (Embase and PsychINFO) using similar terms:

((craving[Title/Abstract] OR relapse[Title/Abstract] OR recurrence[Title/Abstract] OR abstinence[Title/Abstract] OR recovery[Title/Abstract] OR remission[Title/Abstract]) AND ("sex differences"[Title/Abstract] OR "sex difference"[Title/Abstract] OR "gender differences"[Title/Abstract] OR "gender difference"[Title/Abstract])) AND (opioid[Title/Abstract] OR methamphetamine[Title/Abstract] OR estrous cycle[Title/Abstract] OR estradiol[Title/Abstract] OR progesterone[Title/Abstract]). Filters: Other Animals

Additionally, we performed a manual PubMed search (no time limit) of preclinical studies on sex differences in which rodents were trained to self-administer psychostimulant (cocaine and methamphetamine) or opioid (heroin, oxycodone, and fentanyl) drugs, and investigators used different variations of the extinction-reinstatement model, the incubation of drug craving model, and the relapse after forced abstinence model ^1^. We used a search strategy that included different combinations of the terms “sex differences” and “rat” or “mouse” and the following terms: “relapse”, “reinstatement”, “extinction”, “craving”, “cocaine”, “methamphetamine”, “heroin”, “opioids”, “estrous cycle”, “estradiol”, and “progesterone.” We limited our search to studies published in English language and excluded reviews and conference abstracts. We also identified relevant articles that were not identified by the formal literature search by screening the reference lists of the identified articles. Finally, we used the PubMed tools “Similar articles” and “Cited by” to identify additional relevant articles. The articles we identified in the systematic and manual searches were combined and screened independently by CN and YS, in consultation with NEZ in cases of disagreement (**Figure S2**). We used EndNote X9 to collect, de-duplicate, and manage the records.

**Figure S1. A flow chart of the clinical literature search and review**

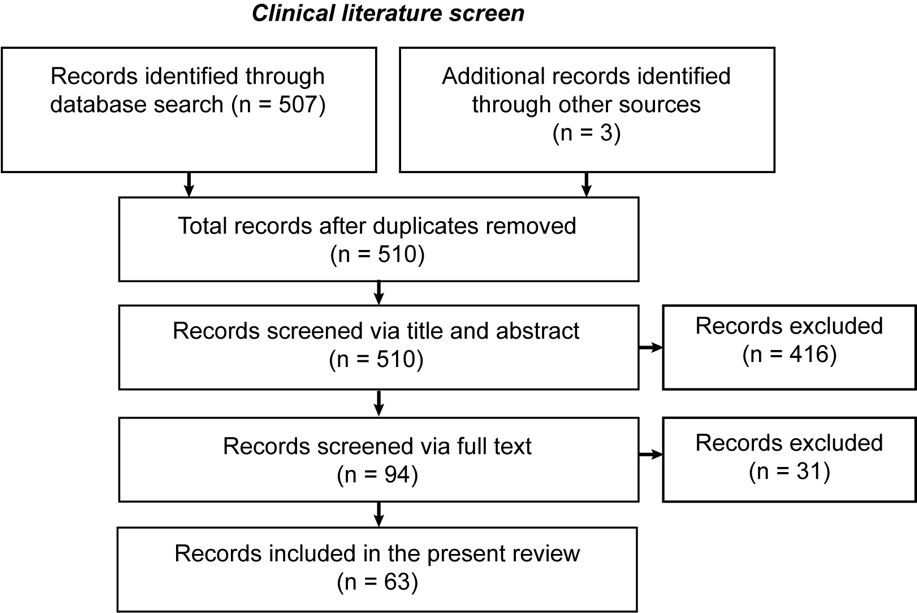

**Figure S2. A flow chart of the preclinical literature search and review**

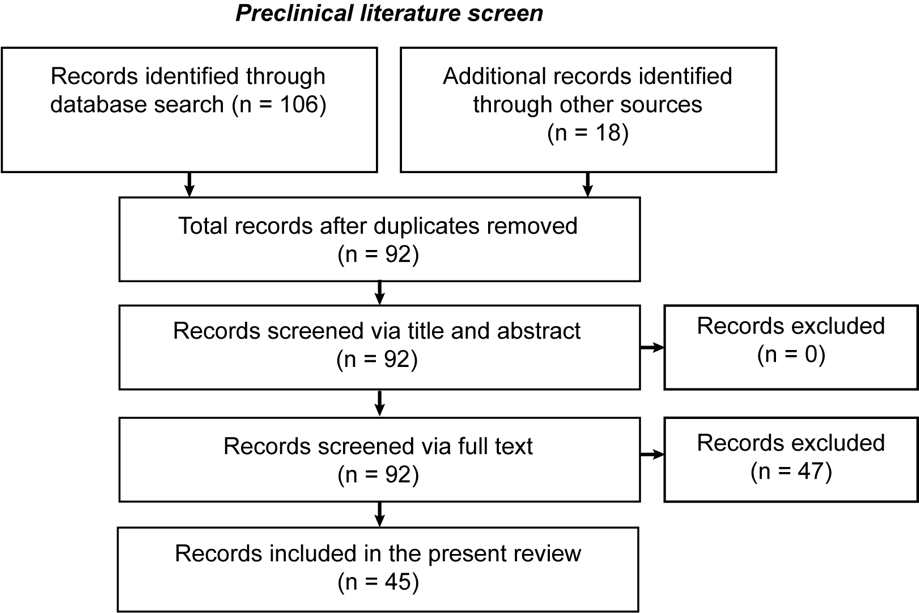

**Additional human neuroimaging studies on sex differences**

Below we describe additional neuroimaging studies that (1) separately analyzed the data within each sex by comparing cue or stress conditions to a neutral cue, or (2) compared, within each sex, brain responses to cocaine cue, stress cue, and neutral cue between cocaine-experienced and drug-naïve control subjects. We provide a summary of these studies in **Table S5**.

Li and Sinha ^7^ used data from the same subjects participated in Li et al. ^8^ study. They investigated stress-induced brain activation and its association with alexithymia (decreased capacity to identify and describe one's own feelings). The main finding was that men showed positive correlation between alexithymia scores (Toronto Alexithymia Scale) and stress-induced activation of the right putamen and middle frontal cortex. In contrast, women showed negative correlations between alexithymia scores and stress-induced activation of the right amygdala, thalamus, putamen, and left frontal and bilateral temporal cortices. Thus, stress-induced brain activity during cocaine abstinence appears to interact with alexithymia in a sex-specific manner. However, the result of the two studies should be interpreted with caution because of the small sample size for an imaging study.

Li et al. ^9^ investigated brain activity using fMRI during cue and stress imagery in 6 women and 5 men who were cocaine abstinent for at least two weeks. There were no sex differences in cue- or stress-induced craving. However, men showed higher activation in the left uncus and right claustrum during cue versus stress trials. No brain regions showed higher activation during stress versus cue trials in men. In contrast, women showed higher activation in the right medial and superior frontal gyri during stress versus cue trials, but no brain regions showed higher activation during cue versus stress trials in women. The results of this study suggest both quantitative and qualitative sex differences in stress- vs. cue-induced brain activity. However, these results should be interpreted with caution because of the low number of subjects.

Potenza et al. ^10^ investigated brain activity using fMRI during cocaine cue and stress cue imagery in inpatient men and women (n=14-16 per sex) who were abstinent from cocaine for at least two weeks. The authors also included a cocaine-naïve control group (n=18 per sex). As previously shown (see above and main text), cocaine and stress cues increased drug craving in both sexes. However, analysis of the brain imaging data within each sex showed differences in the pattern of brain activation during stress and drug cue trials. In women, stress-induced craving in cocaine abstinent subjects was associated with increased activity in amygdala, hippocampus, lateral and medial ventral prefrontal cortices, ventral and dorsal striatum, insula, anterior cingulate, temporal and parietal cortices, and dorsomedial and dorsolateral cortices. In contrast, in men, stress-induced craving was associated with more restricted activation of the striatum, thalamus and temporal cortex. Additionally, in women, cue-induced craving was associated with decreased activation of the posterior cingulate, dorsal frontal, and parietal. In contrast, in men, cue-induced craving was associated with increased activation of the amygdala, hippocampus, ventral and lateral prefrontal cortices, insula, ventral striatum, thalamus, anterior and posterior cingulate, temporal and parietal cortices, and dorsolateral and dorsomedial prefrontal cortices. Finally, in women, cue-induced craving (but not stress-induced craving) was positively correlated with activation of midbrain, hippocampus, ventrolateral prefrontal cortex, temporal cortex, cerebellum and thalamus. In men, cue-induced craving was positively correlated with activation of hippocampus, insula, posterior cingulate, dorsolateral and dorsomedial prefrontal cortices, temporal and parietal cortices, and cerebellum; stress-induced craving was correlated with activation of cerebellum and parietal cortices. No significant effects of the menstrual cycle phases on brain activity were found in this study; however, these results are not conclusive because of the small sample size.

The authors of this comprehensive study interpreted the results to suggest that in cocaine-dependent women corticostriatal-limbic hyperactivity mainly contributes to stress reactivity, while in men corticostriatal-limbic hyperactivity mainly contributes cue reactivity. However, this conclusion should be interpreted with caution for two reasons. First, in both men and women, exposure to the neutral cues also activated corticostriatal-limbic regions. Second, the conclusions are not based on statistical comparisons between men and women but on comparisons within each sex between cocaine-experienced and cocaine-naïve subjects.

**Table S1**. Psychostimulant and opioid craving and relapse: clinical studies. Abbreviations: METH: methamphetamine, M: Male, F: Female, n.s: not significant

| Drug | Active drug use status and n per sex | Measure(s) | Effect | *p* value | Reference |
| --- | --- | --- | --- | --- | --- |
| Psychostimulant | | | | | |
| Cocaïne | | | | | |
| Human laboratory studies | | | | | |
| Cocaine | Cocaine, 10+ h abstinent  F = 5  M = 28 | Cocaine-induced cocaine craving (0.2 mg/kg, i.v.) | F = M | n.s. | ^11^ |
| Cocaine | Cocaine, < 7 d abstinent  F = 9  M = 11 | Cocaine-induced cocaine craving (up to 6 50-mg doses/session, smoked) | F < M | <0.05 | ^12^ |
| Cocaine | Cocaine, 12+ h abstinent  F = 10  M = 11 | Cocaine craving questionnaire | F > M | < 0.05 | ^13^ |
| Cocaine | Cocaine, 7-14 d abstinent  F = 18  M = 24 | Cocaine craving questionnaire | F = M | n.s | ^14^ |
| Cocaine | Cocaine, 7 d abstinent  F = 51  M = 72 | Cocaine craving questionnaire | F = M | n.s | ^15^ |
| Cocaine | Cocaine  F = 10  M = 16 | Cue-induced cocaine craving | F = M | n.s | ^16^ |
| Cocaine | Abstinent, confirmed, outpatient, > 1 mo.  F = 34  M = 47 | Cue-induced cocaine craving | F = M | n.s. | ^17^ |
| Cocaine | Cocaine  F = 12  M = 18 | Cue-induced cocaine craving | F = M | n.s. | ^18^ |
| Cocaine | Abstinent, 7 d inpatient  F = 17  M = 29 | Cue-induced cocaine craving | F = M | n.s. | ^19^ |
| Cocaine | Abstinent, 14 d inpatient  F = 16  M = 14 | Cue-induced cocaine craving | F = M | n.s | ^10^ |
| Cocaine | Abstinent, 14 d inpatient  F = 25  M = 25 | Baseline cocaine craving  Cue-induced cocaine craving | F = M  F = M | n.s  n.s | ^20^ |
| Cocaine | Abstinent, 14 d inpatient  F = 17  M = 23 | Cue-induced cocaine craving | F = M | n.s. | ^21^ |
| Cocaine, alcohol, nicotine | Nicotine, 14-21 d inpatient  F = 13  M = 27 | Cue-induced cocaine craving | F > M | <0.001 | ^22^ |
| Cocaine | Abstinent, unconfirmed, outpatient  F = 38  M = 26 | Cue-induced cocaine craving | F > M | <0.05 | ^23^ |
| Cocaine | Cocaine, 2 d abstinent  F = 25  M = 28 | Cue-induced cocaine craving  Stress-induced cocaine craving | F = M  F > M | n.s  0.0015 | ^24^ |
| Cocaine | Cocaine, 3 d abstinent  F = 21  M = 18 | Stress-induced cocaine craving | F = M | n.s | ^25^ |
| Cocaine | Cocaine, 3 d abstinent  F = 25  M = 28 | Stress-induced cocaine craving | F = M | n.s. | ^26^ |
| Cocaine | Abstinent, 14+ d inpatient  F = 10  M = 17 | Stress-induced cocaine craving | F = M | n.s | ^8^ |
| Cocaine | Cocaine, 3 d abstinent  F = 30  M = 32 | Cue-induced cocaine craving following yohimbine injections (21.6 mg) | F > M | 0.006 | ^27^ |
| Cohort studies | | | | | |
| Cocaine | Abstinent  F = 39  M = 87 | Relapse during treatment (3 mo.) | F < M | <0.0001 | ^28^ |
| Cocaine, Opioids | Opioid agonist treatment  F = 189  M = 134 | Relapse during treatment (3 mo.) | F < M | 0.001 | ^29^ |
| Cocaine | Abstinent  F = 104  M = 350 | Relapse during treatment (6 mo.) | F < M | 0.015 | ^30^ |
| Cocaine | Abstinent  F = 37  M = 64 | Relapse at follow-up (6 mo.) | F < M | <0.02 | ^31^ |
| Cocaine | Abstinent  F = 19  M = 53 | Relapse during treatment (21 days)  Relapse at follow-up (6 mo.) | F = M  F < M | n.s  <0.05 | ^32^ |
| Cocaine, Heroin | Opioid agonist treatment  F = 42  M = 72 | Relapse during treatment (6 mo.)  Cocaine craving during abstinence (6 mo.) | F > M  F = M | 0.0003  n.s | ^33^ |
| Cocaine | Abstinent  F = 34  M = 47 | Relapse at follow-up (9 mo.) | F = M | n.s. | ^17^ |
| Cocaine | Abstinent  F = 77  M = 244 | Relapse at follow-up (12 mo.) | F = M | n.s. | ^34^ |
| Cocaine | Abstinent  F = 10  M = 26 | Relapse at follow-up (12 mo.) | F = M | n.s | ^35^ |
| Cocaine,  Opioids | Opioid agonist treatment  F = 36  M = 80 | Relapse over follow-up period (2 yr.) | F = M | n.s. | ^36^ |
| Methamphetamine | | | | | |
| Human laboratory studies | | | | | |
| METH | METH, 1+ d abstinent  F = 33  M = 10 | cue-induced METH craving | F = M | n.s | ^37^ |
| Cohort studies | | | | | |
| METH | Abstinent  F = 536  M = 329 | METH craving during abstinence (4 mo.) | F = M | n.s | ^38^ |
| METH | Abstinent  F = 225  M = 195 | Relapse during treatment (4 mo.)  Relapse at follow-up (6, 12 mo.) | F > M  F = M | 0.013  n.s | ^39^ |
| METH | METH, regular use  12 mo.:  F = 79  M = 122  5 yr.:  F = 43  M = 59 | Relapse during follow-up (12 mo., 5 yr.) | F < M | 0.005 | ^40^ |
| Cross-sectional studies | | | | | |
| METH | Abstinent  F = 279  M = 1185 | Rate of relapse | F = M | n.s | ^41^ |
| METH | Abstinent  F = 154  M = 196 | Rate of relapse | F = M | n.s. | ^42^ |
| METH | Abstinent  F = 33  M = 65 | Time to relapse | F = M | n.s. | ^43^ |
| Opioids | | | | | |
| Human laboratory Studies | | | | | |
| Heroin | Heroin, inpatient  F = 23  M = 26 | cue-induced heroin craving | F > M | 0.014 | ^44^ |
| Opioids | Opioids  F = 293  M = 599 | opioid craving | F > M | <0.01 | ^45^ |
| Cohort studies | | | | | |
| Opioids | Opioid antagonist treatment  F = 242  M = 223 | Craving during treatment (3 wk.) | F = M | n.s. | ^46^ |
| Opioids | Opioid agonist treatment  F = 47  M = 135 | Stress-induced opioid craving (4 mo.) | F > M | <0.0001 | ^47^ |
| Cocaine, Heroin | Opioid agonist treatment  F = 42  M = 72 | Relapse during treatment (6 mo.)  Heroin craving during abstinence (6 mo.) | F = M  F = M | n.s  n.s | ^33^ |
| Heroin | Residential inpatient treatment  F = 110  M = 150 | Relapse over follow-up period (2 mo.) | F < M | < 0.01 | ^48^ |
| Opioids | Opioid agonist treatment  F = 163  M = 277 | Relapse during treatment (3 mo.) | F = M | n.s. | ^49^ |
| Heroin | Opioid agonist treatment  F = 31  M = 60 | Relapse during treatment (6 mo.) | F > M | 0.027 | ^50^ |
| Opioids | Opioid agonist treatment  F = 117  M = 173 | Relapse during treatment (12 mo.) | F = M | n.s. | ^51^ |
| Heroin | Opioid agonist treatment  F = 54  M = 46 | Relapse during treatment (12 mo.) | F = M | n.s. | ^52^ |
| Heroin | Opioid agonist treatment  F = 215  M = 622 | Relapse at follow-up (12 mo.) | F = M | n.s. | ^53^ |
| Heroin | Opioid agonist treatment  F = 63  M = 148 | Relapse at follow-up (12 mo.) | F < M | No p value | ^54^ |
| Opioids,  Cocaine | Opioid agonist treatment  F = 36  M = 80 | Relapse over follow-up period (2 yr.) | F < M | < 0.001 | ^36^ |
| Heroin | Abstinent, different therapeutic programs and prisons  F = 70  M = 178 | Relapse at follow-up (2 yr.)  Relapse at follow-up (7 yr.) | F < M F = M | 0.05  n.s. | ^55^ |
| Heroin | Heroin, regular use  F = 259  M = 277 | Relapse over follow-up period (8 yr.) | F = M | n.s. | ^56^ |
| Heroin | Abstinent, different therapeutic programs plus a not in treatment group  3 yr.:  F = 151  M = 278  11 yr.:  F = 155  M = 276 | Relapse over follow-up period (3 yr.)  Relapse at follow-up (3 yr.)  Relapse over follow-up period (11 yr.)  Relapse at follow-up (11 yr.) | F < M F = M  F < M F = M | No p values | ^57,58^ |
| Cross-sectional studies | | | | | |
| Heroin | Abstinent, different therapeutic programs and prisons  F = 241  M = 547 | Rate of relapse | F = M | n.s | ^59^ |
| Opioids | Opioid agonist treatment  F = 95  M = 2581 | Rate of relapse | F = M | n.s. | ^60^ |

**Table S2.** Psychostimulant and opioid reinstatement and incubation of craving: preclinical studies**.** Abbreviations: SA: Self-administration, FR: fixed ratio, inf: infusion, METH: methamphetamine, M: Male, F: Female, n.s: not significant

| Drug | SA procedure, n per sex, reinforcement schedule,  unit dose | Relapse test | Effect | *p* value | Reference |
| --- | --- | --- | --- | --- | --- |
| Psychostimulant | | | | | |
| Cocaïne | | | | | |
| Reinstatement | | | | | |
| Cocaine | 2 h/d  F = 8  M = 8  FR1  0.2 mg/kg/inf | Cocaine priming (0.32, 1, 3.2 mg/kg) | F > M (1, 3.2 mg/kg)  F = M (0.32 mg/kg) | <0.05  n.s | ^61^ |
| Cocaine | 14 d, 2 h/d  F = 57  M = 18  FR1  0.4 mg/kg/inf | Cocaine priming (5, 10, 15 mg/kg) | F > M (5 mg/kg)  F = M (10, 15 mg/kg) | <0.05  n.s | ^62^ |
| Cocaine | 14 d, 2 h/d  F = 14  M = 14  FR1  0.5 mg/kg/inf | Cocaine priming (15, 30 mg/kg) | F > M | 0.001 | ^63^ |
| Cocaine | 30 d, 2 h/d  Per dose:  F = 7-9  M = 7-12  FR5  0.25, 0.75 mg/kg/inf | Cocaine priming (10 mg/kg) | F < M | < 0.01 | ^64^ |
| Cocaine | 14 d, 2 h/d  Cocaine:  F = 16  M = 19  Footshock:  F = 6  M = 29  FR4  0.2 mg/ml | Cocaine priming (1.25, 2.5 or 5.0 mg/kg)  Intermittent footshock | F > M (2.5, 5 mg/kg)  F < M | <0.001  <0.01 | ^65^ |
| Cocaine | 30 d, 6 h/d  F = 16  M = 18  FR1  0.4 mg/kg/inf | Cocaine priming (0.2, 0.4, 0.8, 1.6 mg/kg)  Cue | F = M  F = M | n.s  n.s | ^66^ |
| Cocaine | 10 d, 6 h/d  F = 29  M = 22  FR1  0.4 mg/kg/inf | Cocaine priming (10 mg/kg)  Cue  Yohimbine (2.5 mg/kg)  Yohimbine (2.5 mg/kg) + cue | F = M  F = M  F = M  F = M | n.s  n.s  n.s  n.s | ^67^ |
| Cocaine | 10 d, 6 h/d  F = 45  M = 43  FR1  0.4 mg/kg/inf | Cocaine priming (10 mg/kg)  Cue | F = M  F = M | n.s  n.s | ^68^ |
| Cocaine | 10 d, 24 h/d  F = 37  M = 34  FR1  1.5 mg/kg/inf | Cue | F = M | n.s | ^69^ |
| Cocaine | 10 d, 2 h/d  Per dose:  F = 16-18  M = 7-12  FR1  0.25-1 mg/kg/inf | Cue | F = M  F < M (for training dose 0.25 mg/kg) | n.s  <0.01 | ^70^ |
| Cocaine | 10 d, 2 h/d  F = 7  M = 7  FR1  0.6 mg/kg/inf | Cue | F > M | <0.05 | ^71^ |
| Cocaine | 12 d, 2 h/d  F = 32  M = 20  FR1  0.5 mg/kg/inf | Cue | F = M | n.s | ^72^ |
| Cocaine | 13 d, 2 h/d  F = 45  M = 45  FR5  0.5 mg/kg/inf | Cue | F = M | n.s | ^73^ |
| Cocaine | 10/14 d, 2 h/d  F = 47  M = 14  FR1  0.5 mg/kg/inf | Cue + Yohimbine (1.25, 2.5 mg/kg) | F > M | < 0.01 | ^74^ |
| Cocaine | 14 d, 2 h/d  F = 11  M = 8  FR1  0.4 mg/kg/inf | Yohimbine (2.5 mg/kg) | F > M | <0.05 | ^75^ |
| Cocaine | 10 d, 2 h/d  F = 20  M = 22  FR1  0.5 mg/kg/inf | Corticotropin-releasing factor (2 µg) | F = M (all population)  F > M (high responders) | n.s  < 0.05 | ^76^ |
| Cocaine | 10 d, 2 h/d  F = 22  M = 22  FR1  0.5 mg/kg/inf | Footshock (0.5 sec, 0.5 mA) | F > M | < 0.05 | ^77^ |
| Incubation of craving | | | | | |
| Cocaine | 10 d, 2 h/d  Per condition:  F = 14-17  M = 14-16  FR1  0.5 mg/kg/inf | Abstinence day 1, 14, 60 and 180 | F > M | <0.05 | ^78^ |
| Cocaine | 12 d, 8 h/d  Continuous or intermittent:  F = 23-24  M = 25-27  FR1  0.75 mg/kg/inf | Abstinence day 2 and 29 | F > M | <0.001 | ^79^ |
| Cocaine | Non-contingent cocaine-cue pairings,  24 pairings, 0.3 mg/kg/inf; Self-admin., 7d, 2 h/d  Per condition:  F = 4 or 9  M = 9  FR1  0.8 mg/kg/inf | Abstinence day 1 and 30 | Trend: F > M | n.s. | ^80^ |
| Cocaine | 10 d, 3 h x 2/d  F = 6  M = 5  FR1  0.75 mg/kg/inf | Abstinence day 1, 21, 60, 120, 200, 300,400 | F > M | 0.0084 | ^81^ |
| Methamphetamine | | | | | |
| Reinstatement | | | | | |
| METH | 10 d, 2 h/d  F = 9  M = 10  FR1  0.05 mg/kg/inf | METH priming (1 mg/kg) | F > M | <0.01 | ^82^ |
| METH | 7 d, 8 h/d  Per condition:  F = 7-8  M = 7-8  FR1  0.09-0.12 mg/inf | METH priming (1 mg/kg) | F > M | <0.05 | ^83^ |
| METH | 14 d, 1 or 6 h/d  Per condition:  F = 9-10  M = 6-8  FR1  17.5-20 ug/inf | METH priming (0.03, 0.1, 0.3,1 and 3 mg/kg) | F > M (0.3 mg/kg for 1 h SA, 1 mg/kg for 6 h SA) | <0.05 | ^84^ |
| METH | 14 d, 2 h/d  F = 39  M = 22  FR5  17.5-20 ug/inf | METH priming (1 mg/kg)  Cue  Yohimbine (2.5 mg/kg) | F > M  F > M  F > M | <0.05  <0.05  <0.05 | ^85^ |
| METH | 12 d, 2 or 6 h/d  Per condition:  F = 16  M = 16  FR1  0.1 mg/kg/inf | METH priming (0.3, 1.0 mg/kg)  Yohimbine (0.625, 1.25 mg/kg) | F = M  F = M | n.s  n.s | ^86^ |
| METH | 14 d, 2 h/d  F = 20  M = 20  FR1  0.1 mg/kg/inf | METH priming (1 mg/kg)  Cue | F < M  F = M | 0.038  n.s | ^87^ |
| METH | 14 d, 6 h/d  F = 17  M = 22  FR1?  0.05 mg/kg/inf | Cue | F < M | <0.05 | ^88^ |
| METH | 13 d, 2 h/d  F = 27  M = 26  FR5  17.5-20 ug/inf | Cue | F = M | n.s | ^89^ |
| Incubation of craving and relapse after forced abstinence | | | | | |
| METH | 12 d, 6 h/d  Per condition:  F = 10  M = 10-11  FR1  0.1 mg/kg/inf | Abstinence day 1 and 21 | F = M | n.s | ^90^ |
| METH | 20 d, 2x3 h/d  F = 24  M = 24  FR1  0.1 mg/kg/inf | Abstinence day 3 and 30 | F = M | n.s | ^91^ |
| METH | 12 d, 2 or 6 h/d  Per condition:  F = 16  M = 16  FR1  0.1 mg/kg/inf | Abstinence day 2 and 30 | F = M | n.s | ^86^ |
| METH | 14 d, 90 min/d  F = 6  M = 6  FR1  0.08 mg/kg/inf | Forced abstinence 14 days, relapse day 15 | F > M | 0.006 | ^92^ |
| METH | 14 d, 90 min/d  F = 25  M = 16  FR1  0.08 mg/kg/inf | Forced abstinence 14 days, relapse day 15 | F > M | 0.012 | ^93^ |
| Opioids | | | | | |
| Reinstatement | | | | | |
| Heroin | 10 d, 2 h/d  F = 34  M = 30  FR1  0.015 mg/kg/inf | Heroin priming (0.5 mg/kg)  Yohimbine (2.5 mg/kg) | F > M  F > M | <0.05  <0.05 | ^94^ |
| Heroin | 12 d, 3 h/d  F = 7  M = 7  FR4  0.04 mg/inf | Cue | F > M | <0.05 | ^95^ |
| Fentanyl | 10 d, 24 h/d  Per condition:  F = 12-15  M = 10-11  FR1  0.25 µg/kg/inf | Cue | F = M | n.s. | ^96^ |
| Oxycodone | 14 d, 6 h/d  Per condition:  F = 12-13  M = 11-17  FR1  0.05-0.1 mg/kg/inf | Context | F = M | n.s | ^97^ |
| Incubation of craving and relapse after forced abstinence | | | | | |
| Heroin | 12 d, 6 h/d  Per condition:  F = 15-16  M = 11-15  FR1  0.1 mg/kg/inf | Abstinence day 1 and 21 | F = M | n.s | ^90^ |
| Heroin | 12 d, 6 h/d:  Per condition:  F = 16  M = 16-18  FR1  0.1 mg/kg/inf | Abstinence day 1 and 15 | F = M | n.s | ^98^ |
| Fentanyl | 12 d, 6 h/d  F = 20  M = 22  FR1  2.5 ug/kg/inf | Abstinence day 1 and 14 | F = M | n.s | ^99^ |
| Oxycodone | 14 d, 6 h/d  Per condition:  F = 12-25  M = 14-28  FR1  0.1 mg/kg/inf | Abstinence day 1, 15 and 30 | F = M | n.s | ^100^ |

**Table S3.** Role of menstrual cycle in cocaine subjective effects and craving. Abbreviations: n.s: not significant; F: females

| Cocaine status and sample size | Measures | Effect | *p* value | Reference |
| --- | --- | --- | --- | --- |
| Single smoked cocaine dose  F = 21 | Craving  Feel high | Follicular > luteal  Follicular > luteal | 0.019  0.03 | ^101^ |
| Repeated smoked cocaine doses  F = 11 | Subjective positive drug effects, craving | Follicular > luteal | <0.05 | ^102^ |
| Repeated smoked cocaine doses  F = 8 | Subjective positive drug effects | Follicular = luteal | n.s | ^103^ |
| Acute intranasal dose of cocaine  F = 7 | Subjective positive drug effects | Follicular = luteal | n.s | ^104^ |
| Abstinent to smoked cocaine  F = 12 | Craving | Follicular = luteal | n.s | ^105^ |
| Engaged in treatment  F = 16 | Cue-induced craving | Follicular = luteal | n.s | ^10^ |

**Table S4.** Role of estrous cycle in cocaine reinstatement and incubation of craving
Abbreviations: SA: Self-administration, FR: fixed ratio, Inf: infusion, E: estrus, D: diestrus, P: proestrus, NE: non-estrus, n.s: not significant; F: females

| SA procedure, sample size, reinforcement schedule, unit dose | Relapse tests | Effect | *p* value | References |
| --- | --- | --- | --- | --- |
| Cocaine | | | | |
| Reinstatement | | | | |
| 10 d, 2 h/d  F = 73  FR1  0.5 mg/kg/inf | Cocaine priming (5, 10 mg/kg) | E > D, P, male (10 mg/kg) | <0.05 | ^106^ |
| 10 d, 2 h/d  F = 30  FR1  0.5 mg/kg/inf | Cocaine priming (5, 10 mg/kg) | E > D, P (10 mg/kg) | <0.05 | ^107^ |
| 10 d, 2 h/d  F = 61  FR  0.5 mg/kg/inf | Cocaine priming (10 mg/kg) | E > NE, male | <0.05 | ^78^ |
| 10 d, 2 h/d  F = 60  FR1  0.5 mg/kg/inf | Cocaine priming (10 mg/kg) | E > D, P | <0.05 | ^108^ |
| 10 d, 2 h/d  Per dose:  F = 16-18  FR1  0.25-1 mg/kg/inf | Cue | E < NE (only for SA training with 0.25 mg/kg) | <0.01 | ^70^ |
| 10 d, 24 h/d  F = 37  FR1  1.5 mg/kg/inf | Cue | E = NE | n.s | ^69^ |
| 12 d, 2 h/d  F = 32  FR1  0.5 mg/kg/inf | Cue | E = NE | n.s | ^72^ |
| 10/14 d, 2 h/d  F = 47  FR1  0.5 mg/kg/inf | Cue + Yohimbine (1.25, 2.5 mg/kg) | D, E < P | < 0.01 | ^74^ |
| Incubation of craving and relapse after forced abstinence | | | | |
| 10 d, 2 h/d  F = 61  FR  0.5 mg/kg/inf | Abstinence day 1, 14, 60 and 180 | E > NE, male (day 1, 60 and 180) | <0.05 | ^78^ |
| 12 d, 8 h/d  Continuous or intermittent:  F = 23-24  FR1  0.75 mg/kg/inf | Abstinence day 29 | E > NE | <0.05 | ^79^ |
| Methamphetamine | | | | |
| Reinstatement | | | | |
| 14 d, 2 h/d  F = 39  FR5  17.5-20 ug/inf | Meth priming (1 mg/kg) | E = D, P | n.s | ^85^ |
| Opioids | | | | |
| Reinstatement | | | | |
| 10 d, 24 h/d  Per condition:  F = 11-16  FR1  0.25 µg/kg/inf | Cue | E > NE | <0.05 | ^96^ |

**Table S5.** Human neuroimaging studies on cocaine cue- and stress-induced brain activity. Abbreviations: F: female, M: male, fMRI: functional magnetic resonance imaging, PET: positron emission tomography, ^18^FDG: 2-[18F]-fluoro-2-deoxy-d-glucose, PFC: prefrontal cortex, vmPFC: ventromedial prefrontal cortex, PAG: periaqueductal gray, n.s: not significant

| Drug | Active drug use status, N per sex, imaging technique | Drug craving | Brain regions | Effects | *p* value | Reference |
| --- | --- | --- | --- | --- | --- | --- |
| Cue-induced craving | | | | | | |
| Direct comparison between males and females | | | | | | |
| Cocaine | Abstinent, ≥ 3 d  Cocaine-dependent  F = 24  M = 43  fMRI | F = M, n.s. | Dorsomedial PFC  Amygdala | F = M  F = M | n.s.  n.s. | ^109^ |
| Cocaine | Abstinent, 1-14 d  Cocaine-dependent  F = 8  M = 8  PET ([^15^O]H2O) | F = M  n.s. | Amygdala  Insula  Postcentral gyrus  Caudate nucleus  Precentral gyrus  Middle frontal gyrus  Posterior cingulate gyrus | Activation:  F < M  F < M  F < M  F < M  F > M  F > M  F > M | <0.005 | ^110^ |
| Cocaine | Active cocaine users  F = 10  M = 16  PET (^18^FDG) | F = M  n.s. | Frontal cortex  Anterior cingulate cortex  Posterior cingulate cortex  Inferior parietal  Dorsomedial thalamus | Inhibition:  F < M  F < M  F < M  F < M  F < M | <0.001 | ^16^ |
| Cocaine | Abstinent, 7-10 d  Cocaine-dependent  F = 10  M = 42  fMRI | F = M  n.s. | PAG  PAG-vmPFC connectivity | F = M  F = M | n.s.  n.s. | ^111^ |
| Cocaine | Cocaine-dependent  F = 12  M = 18  Controls:  F = 22  M = 23  fMRI | F = M, n.s. | Dorsomedial PFC,  Superior frontal gyrus | Activation:  F < M  F < M | <0.05  <0.05 | ^18^ |
| Indirect comparison between males and females | | | | | | |
| Cocaine | Abstinent, ≥ 14 days  Cocaine-dependent:  F = 16  M = 14  Controls:  F = 18  M = 18  fMRI | F = M,  n.s. | F:  Posterior cingulate,  Dorsal frontal cortex,  Parietal cortex  Midbrain,  Hippocampus,  Ventrolateral PFC,  Temporal cortex,  Cerebellum,  Thalamus | Activation:  Cocaine F < control F  Positively correlated with craving | <0.05  <0.05 | ^10^ |
|  |  |  | M:  Amygdala,  Hippocampus,  Ventral PFC,  Lateral PFC,  Insula,  Ventral striatum,  Thalamus,  Anterior cingulate,  Posterior cingulate,  Temporal cortex,  Parietal cortex,  Dorsolateral PFC,  Dorsomedial PFC  Hippocampus,  Insula,  Posterior cingulate, Dorsolateral PFC,  Dorsomedial PFC,  Temporal cortex,  Parietal cortex,  Cerebellum | Activation:  Cocaine M <  control M  Positively correlated with craving | <0.05  <0.05 |  |
| Stress-induced craving | | | | | | |
| Direct comparison between males and females | | | | | | |
| Cocaine | Abstinent, 14-21 d  Cocaine-dependent  F = 10  M = 17  fMRI | F = M  n.s. | Frontolimbic areas  Anterior cingulate cortex  Posterior cingulate cortex  Insula  Dorsolateral PFC  Medial PFC  Inferior frontal cortex | Activation :  F > M  F > M  F > M  F > M  F > M  F > M  F > M | <0.01 | ^8^ |
| Indirect comparisons between males and females | | | | | | |
| Cocaine | Abstinent, ≥ 14 d  Cocaine-dependent:  F = 16  M = 14  Controls:  F = 18  M = 18  fMRI | F = M, n.s. | F:  Amygdala,  Hippocampus,  Lat. ventral PFC,  Med. ventral PFC  Ventral striatum,  Dorsal striatum,  Insula,  Anterior cingulate,  Temporal cortex,  Parietal cortex,  Dorsomedial cortex,  Dorsolateral cortex | Cocaine F > control F | <0.05 | ^10^ |
|  |  |  | M:  Striatum,  Thalamus,  Temporal cortex  Cerebellum  Parietal cortex | Cocaine M > control M  Positively correlated with craving | <0.05  <0.05 |  |
| Cocaine | Abstinent, ≥ 14 d  Cocaine-dependent  F = 10  M = 17  fMRI | F = M, n.s. | F :  Amygdala  Thalamus  Putamen  Frontal cortex  Temporal cortex | Activation negatively correlated with alexithymia score | <0.001 | ^7^ |
|  |  |  | M :  Putamen  Middle frontal cortex | Activation positively correlated with alexithymia score | <0.001 |  |
| Cue vs. stress-induced craving | | | | | | |
| Indirect comparisons between males and females | | | | | | |
| Cocaine | Abstinent, ≥ 14 d  Cocaine-dependent  F = 6  M = 5  fMRI | F = M, n.s. | F :  Medial frontal gyrus  Superior frontal gyrus | Activation:  Stress > drug cue | <0.01 | ^9^ |
|  |  |  | M :  Uncus  Claustrum | Activation:  Drug cue > stress | <0.01 |  |

* For full statistical reporting of the imaging data, see the original papers.
